# Automated Ischemic Stroke Lesion Detection On Non-Contrast Brain CT: A Large-Scale Clinical Feasibility Test

**DOI:** 10.1101/2025.06.04.25328996

**Authors:** JoonNyung Heo, Wi-Sun Ryu, Jong-Won Chung, Chi Kyung Kim, Joon-Tae Kim, Myungjae Lee, Dongmin Kim, Leonard Sunwoo, Johanna M. Ospel, Nishita Singh, Hee-Joon Bae, Beom Joon Kim

**Affiliations:** Department of Neurology, Yonsei University College of Medicine, Seoul, Korea; Artificial Intelligence Research Center, JLK Inc, Seoul, Republic of Korea; Department of Neurology, Samsung Medical Center, Sungkyunkwan University College of Medicine, Seoul, Republic of Korea; Department of Neurology, Korea University Guro Hospital and Korea University College of Medicine, Seoul, Korea; Department of Neurology, Chonnam National University Hospital, Chonnam National University Medical School, Gwangju, Korea; Department of Radiology, Seoul National University Bundang Hospital, Seoul National University College of Medicine, Seongnam, Republic of Korea; Department of Diagnostic Imaging, Foothills Medical Center, University of Calgary, Calgary, Alberta, Canada; Department of Clinical Neurosciences, Foothills Medical Center, University of Calgary, Calgary, Alberta, Canada; Department of Neurology, Faculty of Medicine, University of Manitoba, Winnipeg, Canada; Department of Neurology, Seoul National University Bundang Hospital, Seongnam, Republic of Korea

## Abstract

**Background:** Non-contrast CT (NCCT) is widely used imaging modality for acute stroke imaging but often fails to detect subtle early ischemic changes. Such underestimation can lead clinicians to overlook tissue-level information. This study aimed to develop and externally validate automated software for detecting ischemic lesions on NCCT and to assess its clinical feasibility in stroke patients undergoing endovascular thrombectomy.

**Methods:** In this retrospective, multicenter cohort study (May 2011–April 2024), a modified 3D U-Net model was trained using paired NCCT and diffusion-weighted imaging (DWI) data from 2,214 patients with acute ischemic stroke. External validation was performed in 458 subjects. Clinical feasibility was assessed in 603 endovascular thrombectomy-treated patients with complete recanalization. Model outputs were compared against expert-annotated DWI lesions for sensitivity, specificity, and volumetric correlation. Clinical endpoints included follow-up DWI lesion volumes, hemorrhagic transformation, and 3-month modified Rankin Scale outcomes.

**Results:** A total of 458 subjects were evaluated for external validation (mean age, 64 years ± 16; 265 men). The model achieved 75.3% sensitivity (95% CI, 70.9–79.9%) and 79.1% specificity (95% CI, 77.1–81.3%). In the feasibility cohort (n = 603; mean age, 69 years ± 13; 362 men), NCCT- derived lesion volumes correlated with follow-up DWI volumes (ρ = 0.60, P < .001). Lesions >50 mL were associated with reduced favorable outcomes (17.3% [26/150] vs 54.2% [246/453], P < .001) and higher hemorrhagic transformation rates (66.0% [99/150] vs 46.3% [210/453], P < .001). Radiomics features improved hemorrhagic transformation prediction beyond clinical variables alone (area under the receiver operating characteristic curve, 0.833 vs 0.626; P = .003).

**Conclusions:** The automated NCCT-based lesion detection model demonstrated reliable diagnostic performance and provided clinically relevant prognostic information in endovascular thrombectomy-treated stroke patients.

## Introduction

Non-contrast computed tomography (NCCT) is the most widely accessible imaging modality for acute stroke worldwide due to its accessibility and utility in rapidly ruling out hemorrhagic stroke.^1^ However, hypodense changes indicative of acute ischemia can be subtle, leading to suboptimal sensitivity.^2^ Although semiquantitative scores are routinely used to communicate the extent of ischemic changes, their inter-rater reliability varies considerably.^3^ Likewise, manual segmentation of early ischemic changes on NCCT often yields low agreement.^4^

Despite these limitations, the initial NCCT scan contains useful tissue-level information such as extent and severity of ischemia, which has not been thoroughly utilized in clinical practice.^5^ Greater emphasis on detecting and quantifying ischemic changes could guide treatment decisions, especially for time-sensitive interventions such as intravenous thrombolysis and endovascular treatment (EVT).

We aimed to develop an automated software model to detect acute ischemic lesions on NCCT. The model’s training and validation used concomitant diffusion-weighted imaging (DWI) with expert ratings as a reference standard, given DWI’s high sensitivity for acute infarction. We further tested the software’s clinical feasibility in a separate cohort of patients with large vessel occlusion (LVO) undergoing EVT with complete recanalization, correlating NCCT-derived lesion volumes and radiomics features with subsequent DWI and clinical outcomes.

## Methods

The study conformed to the Standards for Reporting of Diagnostic Accuracy Studies guidelines for diagnostic accuracy research.^6^

### Acute Ischemic Lesion Detection On NCCT: Model Development and Validation

We retrospectively collected data from six stroke centers in South Korea between 2011 and 2015, including 2,398 ischemic stroke patients with paired NCCT and DWI scans acquired within 3 hours to minimize ischemic lesion evolution. Five expert neurologists each with over 10 years of clinical experience, manually annotated ischemic lesions visible on NCCT while referring to DWI / apparent diffusion coefficient images for confirmation. Experts’ annotation agreements were assessed using volumetric similarity indices and absolute volume difference, with ground truth defined by consensus of more than 2 experts. The labelled 2,398 ischemic stroke patients were randomly categorized into 2,214 cases for training / internal validation cohort and 184 cases for external validation cohort with 274 non-stroke individuals.

A modified 3D U-Net was trained to detect hypoattenuated lesions on NCCT. Expert consensus masks served as ground truth, and any lesion <0.5 mL on either ground-truth or model’s prediction was excluded from the final performance analysis, based on the premise of NCCT’s limited spatial resolution. Model performance was evaluated in the external validation cohort of 732 subjects (458 ischemic stroke patients and 274 non-stroke individuals) without any overlap with the training cohort, yielding sensitivity, specificity, positive predictive value, negative predictive value, and volumetric agreement of Dice similarity coefficient.

Full details regarding patient selection, image preprocessing, model architecture, and evaluation metrics are available in eMethods.

### Automated Ischemic Lesion Detection On NCCT: Clinical Feasibility Testing

#### Patient Selection

For the clinical feasibility assessment, we included 603 consecutive patients who underwent EVT for anterior-circulation LVO and achieved complete recanalization (Thrombolysis in Cerebral Infarction 3). These patients were collected through a multicenter prospective stroke registry of South Korea, the Clinical Research Collaboration for Stroke of Korea ^7,8^, and none of the subjects overlapped with those included for the model development, internal or external validation cohort. All the patients had baseline NCCT, CT perfusion (CTP) images and follow-up DWI scans. Individuals lacking key imaging data, had EVT beyond 24 hours from onset, failing to achieve complete recanalization were excluded.

#### Imaging Analysis

NCCT-based lesion volumes were derived via the automated lesion detection software model. The volume data was compared with those of 1) early post-EVT DWIs, 2) delayed post-EVT DWIs, and 3) baseline CTP before EVT. Early DWIs were defined as scans obtained within 24 hours of NCCT, and delayed DWIs as those acquired between 24 to 168 hours. Infarct volumes on DWIs were calculated using validated software (JLK-DWI, JLK Inc., Republic of Korea), with review by a vascular neurologist (W-S.R.).^9^ Baseline CTP-derived core volumes were obtained using a relative cerebral blood flow <30% threshold, using a validated software (JLK-CTP, JLK Inc., Republic of Korea).^10,11^ Hemorrhagic transformation (HT) on follow-up NCCT, gradient echo MR, or susceptibility-weighted imaging was assessed based on the European Cooperative Acute Stroke Study II criteria.^12^ Radiomic features of the ischemic lesion were extracted from the NCCT lesions. Machine learning models were developed and validated with the radiomic features to predict HT and functional recovery. Detailed methodology is described in eMethods.

### Statistical analysis

Baseline characteristics were compared using ANOVA or Kruskal-Wallis test for continuous variables and chi-square test or Fisher exact test for categorical variables as appropriate. For external validation, volumetric similarity, Dice similarity coefficient, sensitivity, specificity, positive predictive value and negative predictive value of the final ensemble model were evaluated. Spearman’s correlation was used to compare among ischemic lesion volume on the baseline NCCT, ischemic core from CTP, and follow-up DWI, considering for skewed distribution of data.

Categorized ischemic core volumes on NCCT were analyzed for associations with 3-month outcomes and HT. The volume categories were pre-specified as <50 mL vs. ≥50 mL and a more granular stratification as 0-<5mL, 5-<10mL, 10-<20mL, 20-<30mL, 30-<40mL, 40-<50mL, and ≥50-mL. A logistic regression model, adjusted for clinical covariates (age, sex, onset-to-NCCT scan time, NCCT-to-arterial access time, EVT procedure time, intravenous thrombolysis, previous stroke, coronary artery disease, hypertension, diabetes, hyperlipidemia, smoking, and atrial fibrillation) was used to evaluate the association between baseline NCCT core volume and clinical outcomes, with sensitivity analyses comparing volumes from ischemic lesion on NCCT and ischemic core from CTP. Detailed statistical analysis is presented in eMethods.

### Standard Protocol Approvals, Registrations, and Patient Consents

Institutional Review Board at participating centers approved the retrospective analysis and waived additional informed consent requirements due to de-identified data (institutional review board approval # B-2102-667-106).

### Data Availability

The data that support the findings of this study are available upon reasonable request of qualified researchers from the corresponding author. The data are not publicly available due to Korean regulation restrictions and privacy protection.

## Results

### Development and Validation of Automated Acute Ischemic Lesion Detection Software

A total of 2,398 ischemic stroke patients were included for training, internal validation, and external validation (Figure 1A). An additional 274 individuals who presented with stroke-like symptoms but confirmed not to have ischemic lesions on DWI were included as negative controls, yielding 458 subjects in the external validation cohort. The mean ages in the training, internal, and external validation groups were 69.0±12.2, 68.5±12.1, and 64.1±15.5 years, respectively (Table S1). Within the external validation cohort, 89 patients (19.4%) had DWI lesions >0.5 mL, with a median annotated volume of 5.76 mL (interquartile range, IQR: 1.96–20.0 mL). Expert volumetric correlations among the five raters ranged from 0.748 to 0.861, while the absolute volume differences between 2.20 and 6.09 mL (Table S2).

**Figure 1.**
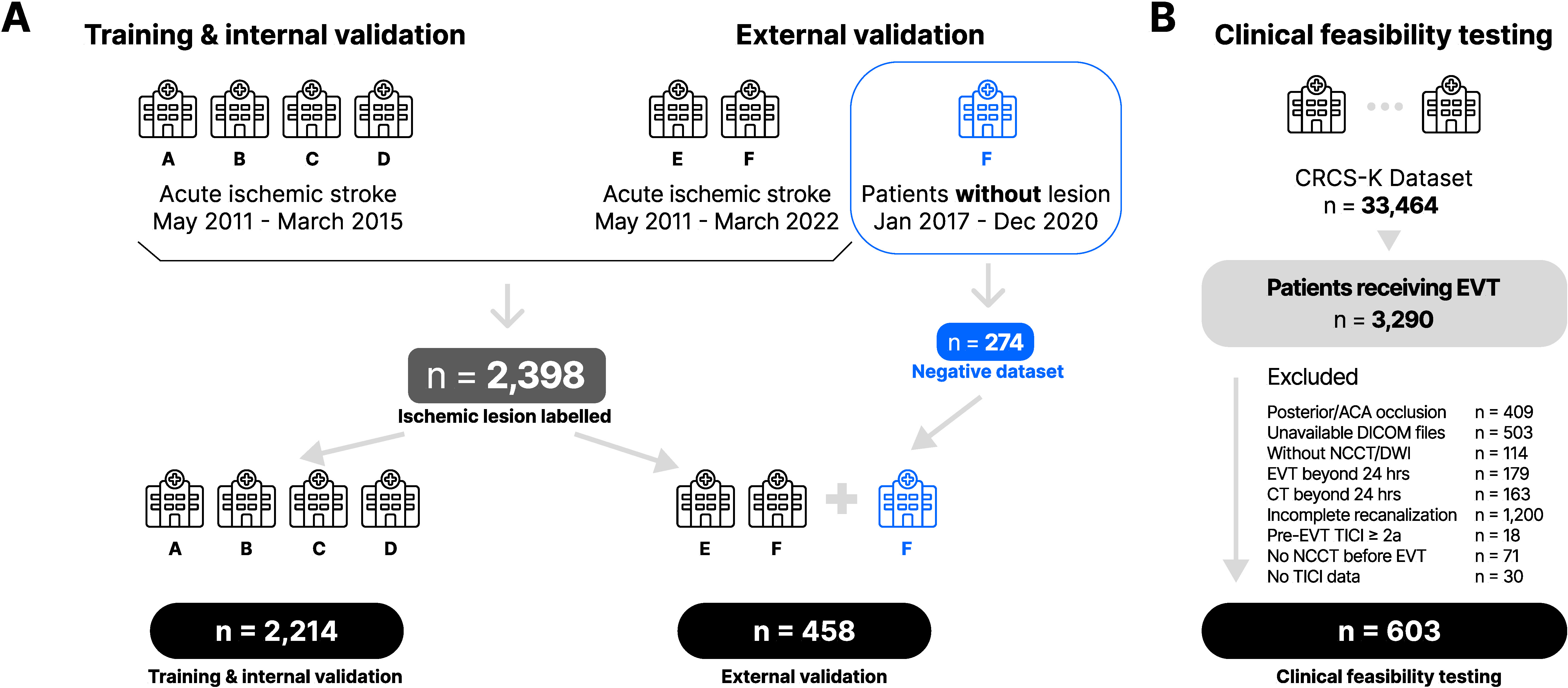
Flowchart for all datasets. NCCT, non-contrast CT; DWI, diffusion-weighted imaging; EVT, endovascular thrombectomy; ACA, anterior cerebral artery; DICOM, Digital Imaging and Communications in Medicine; TICI, thrombolysis in cerebral infarction.

For the external validation cohort of 458 individuals, the automated detection model demonstrated a sensitivity of 75.3% (95% CI: 70.9–79.9%), specificity of 79.1% (77.1–81.3%), a positive predictive value of 49.3% (95% CI: 45.4–53.2%), and a negative predictive value of 95.4% (95% CI: 94.3–96.7%). Notably, the software’s sensitivity improved with increasing lesion volumes, ranging from 53.3% for volumes of 0.5–1 mL to 94.7% for volumes of >30 mL (Table S3). The sensitivity was higher for cases with a single lesion (91.9%) than with multiple lesions (63.5%, Table S4). The predicted lesion volumes demonstrated a robust correlation with the ground truth (Spearman’s ρ = 0.776 (*P* <0.001, Figure S1).

### Clinical Feasibility Testing of Ischemic Lesion Detected from The Automated Software

A total of 603 patients were included to test the clinical feasibility of the software. In the cohort, the mean age was 71.5 (standard deviation 12.8), and 56.4% were male (1B, Table 1). Median National Institutes of Health Stroke Scale score was 14 (IQR, 10–19). The median time from stroke onset to NCCT scan was 152 minutes (IQR 76 – 394).

**Table 1.**
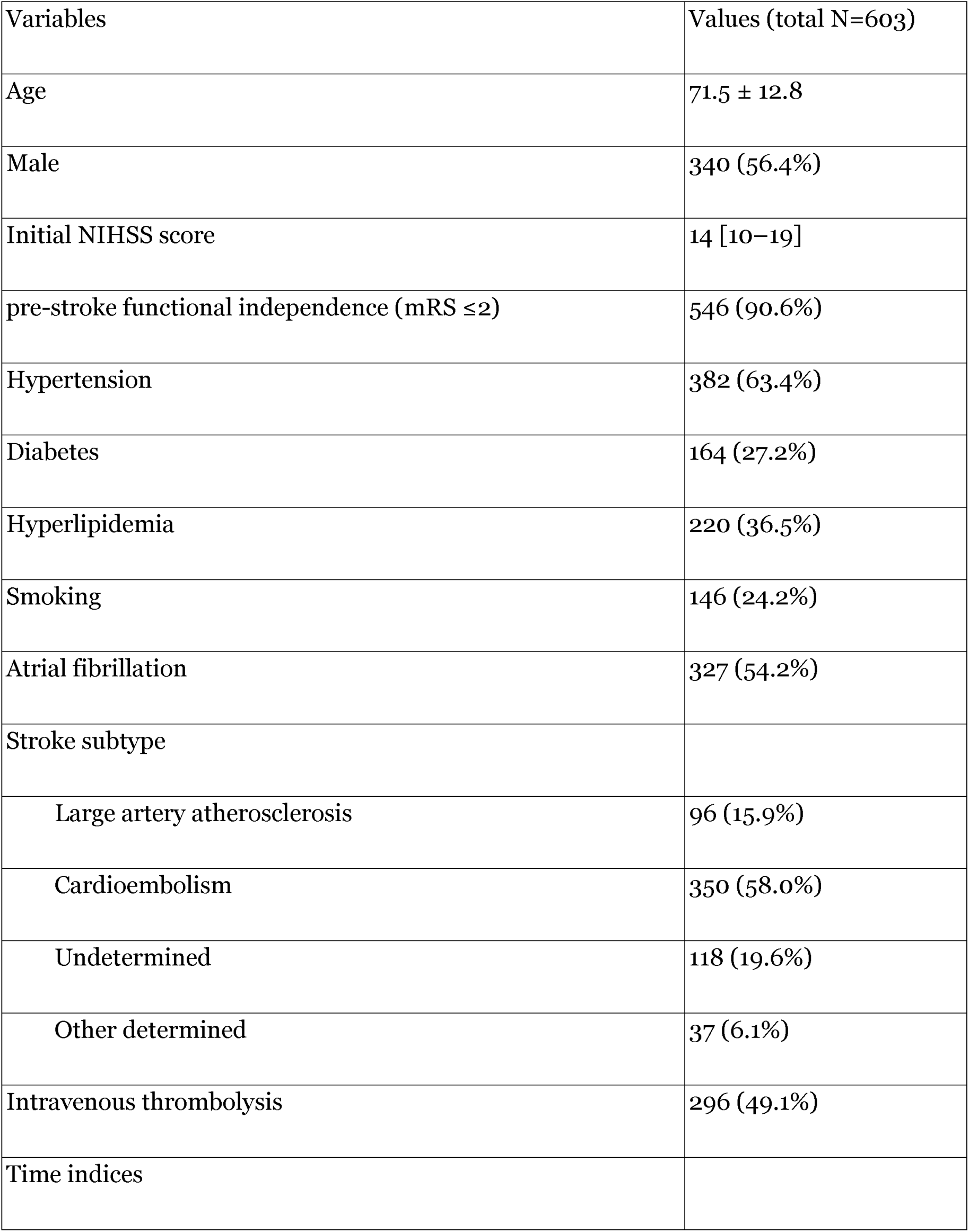
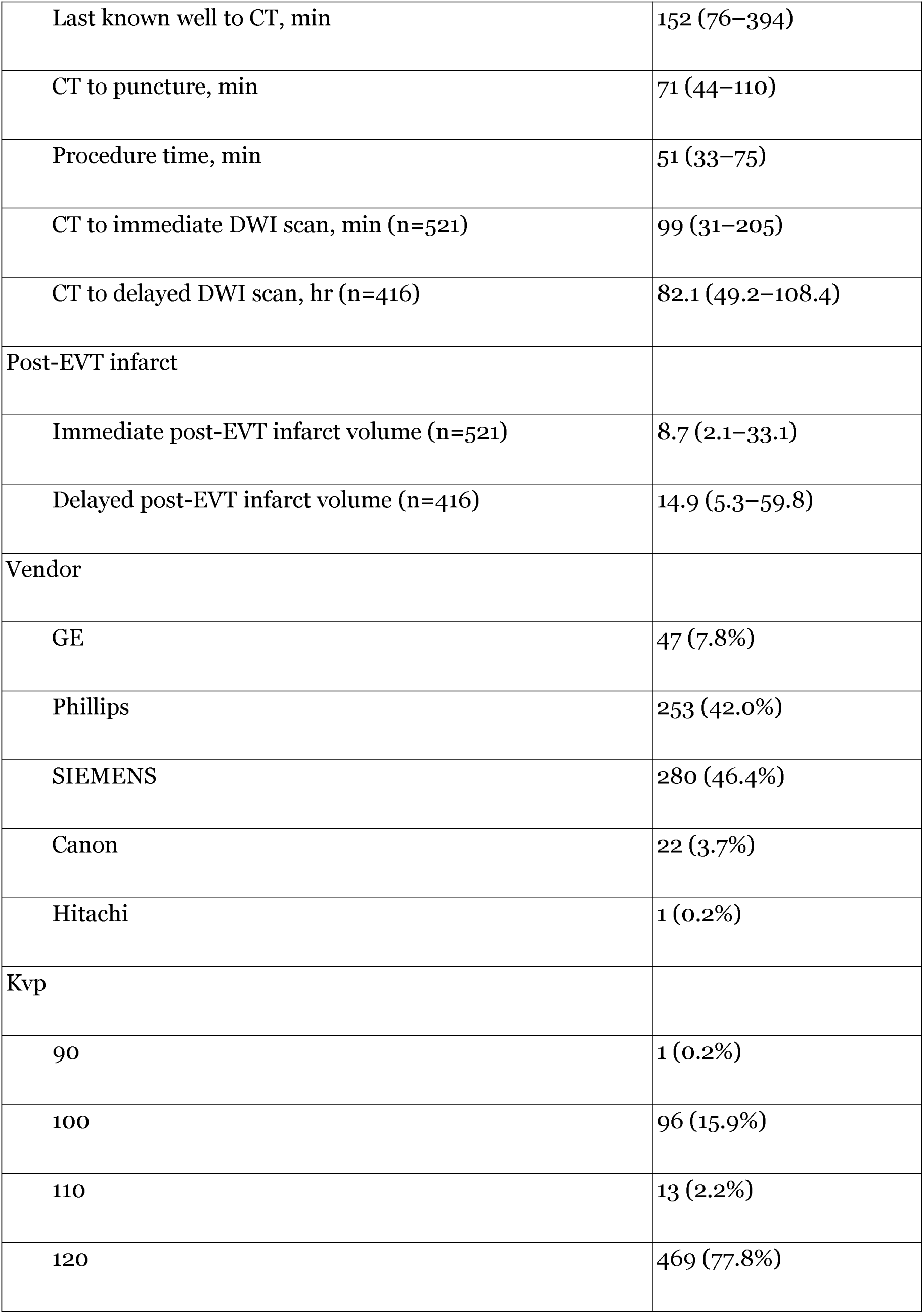
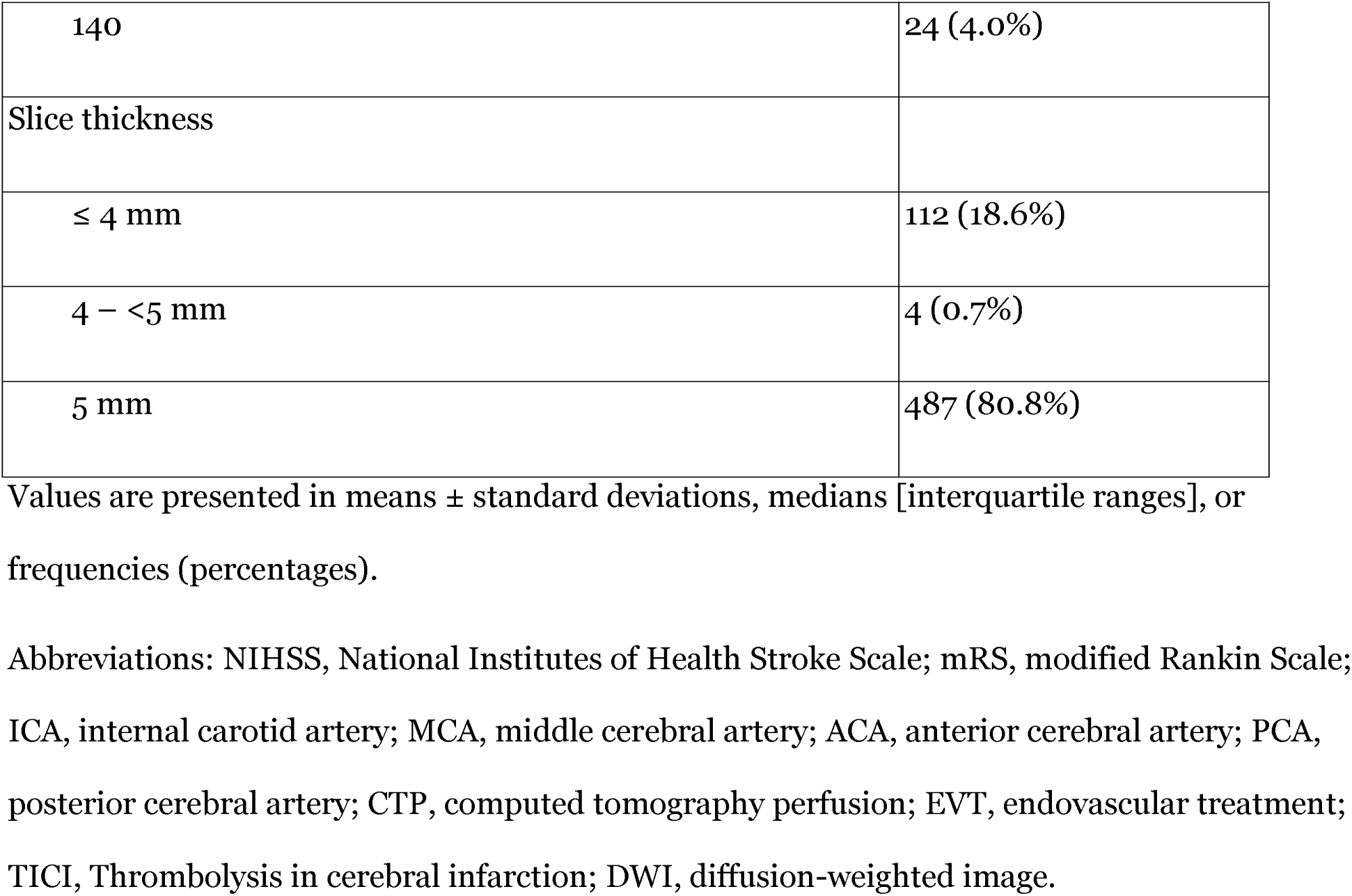
Characteristics of 603 Subjects for Clinical Feasibility Testing of the Model.

#### Comparison of Ischemic Volume on Non-contrast CT with Diffusion Weighted and CT Perfusion Imaging on the Clinical Feasibility Testing Dataset

In the feasibility cohort, the estimated ischemic core volume on NCCT demonstrated a significant correlation with infarct volume on follow-up DWI (Spearman’s ρ = 0.60, *P* <0.001; Figure 2A). The ischemic core volume on CTP was also correlated with early DWI (ρ = 0.50, *P* <0.001; Figure 2B). Overall, NCCT tended to underestimate early follow-up DWI lesion size, whereas CTP tended to overestimate it.

**Figure 2.**
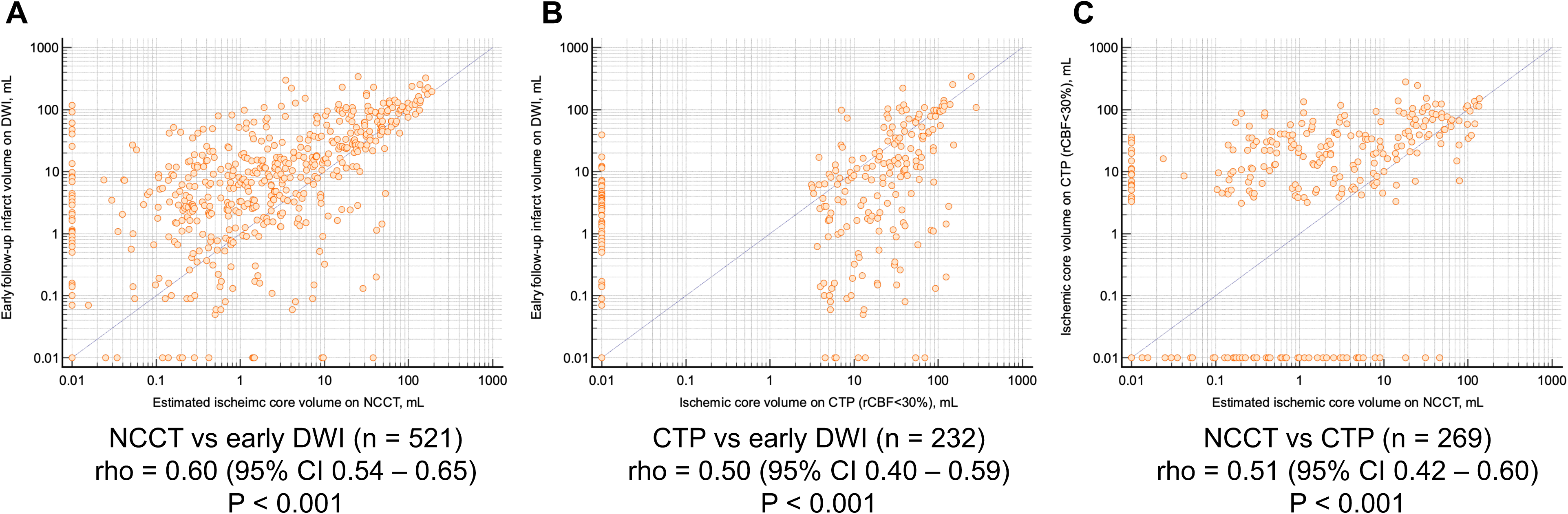
Scatter plots with log-scale between (A) estimated ischemic core non-contrast CT and early follow-up diffusion-weighted imaging infarct volume, (B) ischemic core on CT perfusion and early follow-up diffusion-weighted imaging infarct volume, and (C) estimated ischemic core non-contrast CT and ischemic core on CT perfusion. NCCT, non-contrast CT; DWI, diffusion-weighted imaging; CTP, CT perfusion.

In patients scanned within 180 minutes, both ischemic lesions on NCCT and CTP showed similar correlations with follow-up DWI (ρ = 0.50 vs. 0.51; *P* = 0.89; Figure S2). Beyond 180 minutes, however, NCCT demonstrated a stronger correlation with follow-up DWI than CTP (ρ = 0.74 vs. 0.55; *P* = 0.01; Figure S3). Subgroup analyses by hourly increments further indicated that the correlation between NCCT lesion volumes and DWI infarct size increased over time, surpassing that of CTP after approximately two hours (Table S5). By contrast, when NCCT- and CTP- derived ischemic lesion volumes were compared against late follow-up DWI infarct volume, CTP–based ischemic core demonstrated a stronger correlation than the NCCT (Figure S4).

#### Outcome Prediction with Ischemic Volume on Non-contrast CT

Patients with ischemic lesions >50 mL on NCCT showed favorable outcomes in 17.3%, compared to 54.2% for the patients with lesions ≤50 mL. (Figure 3A). Of the patients with ischemic core lesions >50 mL on CTP, 35.7% showed a favorable outcome (Figure 3B). In the subgroup of patients with NCCT lesion volumes between 0 and 5 mL, more than 60% attained favorable outcomes (Figure S5). A similar trend was observed for volumes up to 40 mL, beyond which the proportion of favorable outcomes declined considerably: only 20% of patients with lesions >40 mL showed favorable recovery. Multivariable analyses also demonstrated a stepwise relationship between larger NCCT lesion volumes and unfavorable outcomes (Table S6), consistent with restricted spline curves (Figure S6).

**Figure 3.**
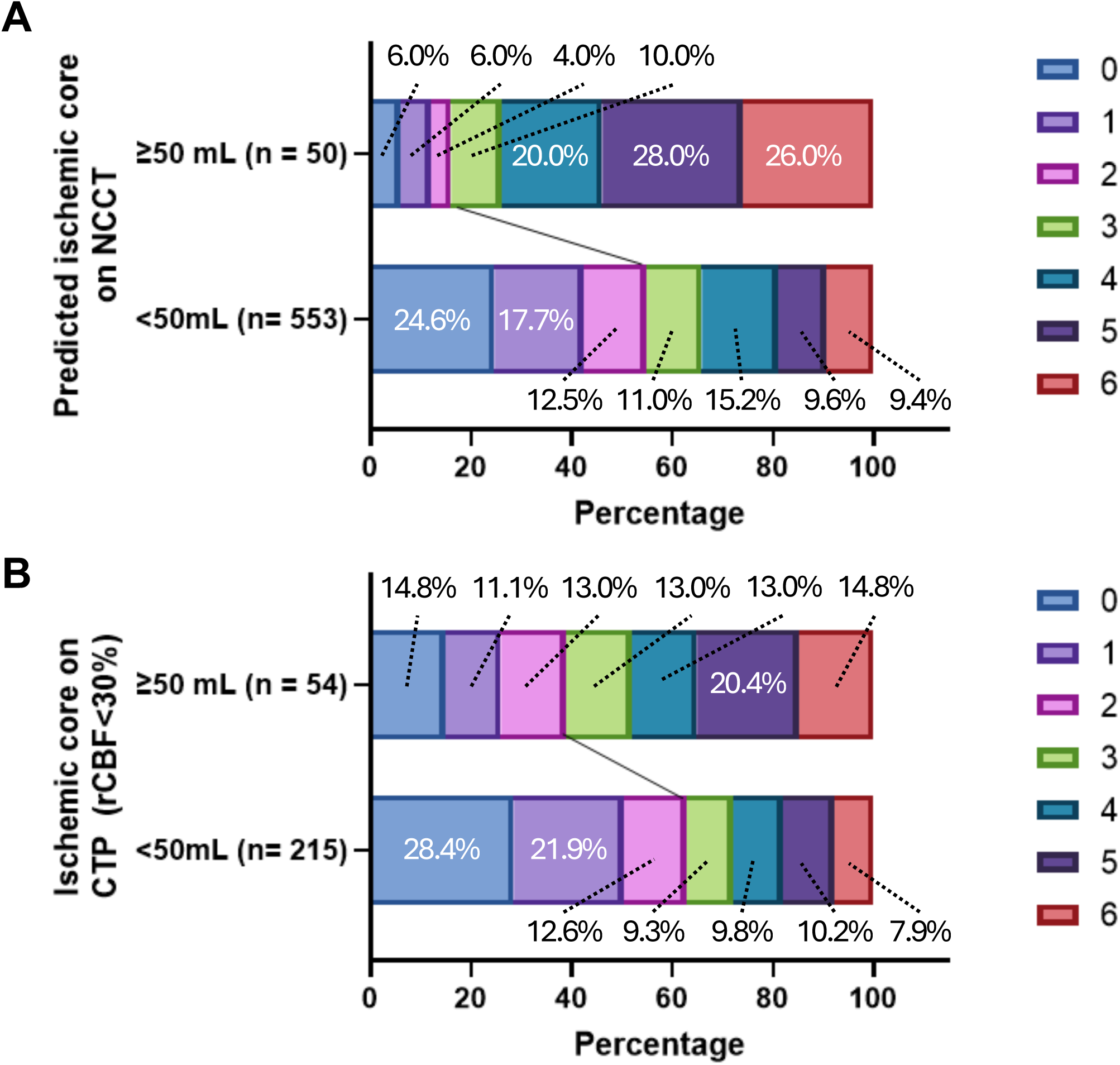
3-month modified Rankin Scale score stratified by estimated ischemic core on (A) non-contrast CT or (B) CT perfusion, ≥ 50 mL versus < 50 mL NCCT, non-contrast CT; CTP, CT perfusion; rCBF, regional cerebral blood flow.

HT occurred in 66.0% of patients with NCCT lesion volumes >50 mL, versus 46.3% for those with ≤50 mL (AUC, 0.805, *P* = 0.04; Figure S7A). Lesions exceeding 5 mL were also linked with a higher risk of HT than lesions of 0–5 mL (Figure S7B). Parenchymal hemorrhage occurred in 40% of patients with lesions >40 mL, a significantly greater proportion than in those with smaller lesions (*P* <0.001). Representative cases are shown in Figure 4.

**Figure 4.**
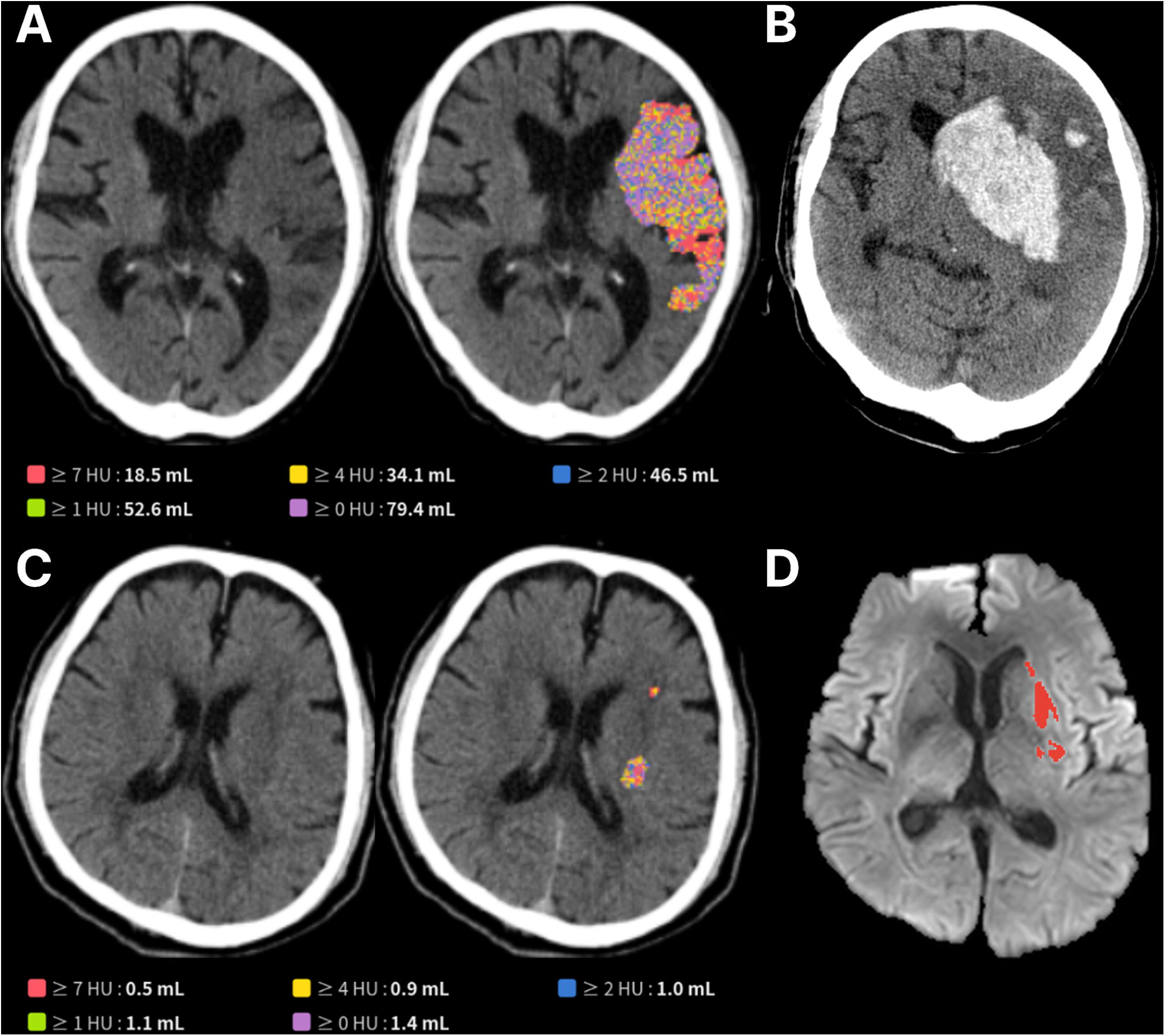
Representative cases showing (A) a large infarct core (79.4 mL) identified on non-contrast CT and (B) subsequently developed parenchymal hematoma type II hemorrhagic transformation while in (C), a small infarct core (1.4 mL) did not result in hemorrhagic transformation (D) on delayed magnetic resonance imaging.

#### Radiomics Features Automatically Extracted from the Ischemic Lesions

First-order radiomics features were extracted from the automatically segmented NCCT ischemic lesions. Energy measures showed increased upper range values and spread in groups with larger lesion volumes (Figure S8). In those with larger lesion sizes, the maximum Hounsfield unit values were generally elevated, while the minimum Hounsfield unit values were shifted toward lower levels. Median Hounsfield unit values were also higher in larger lesions, accompanied by higher values in range and root mean squared. No clear patterns differentiated patients according to the time from symptom onset.

#### Outcome Prediction with Automatically Extracted Radiomics Features

Among the extracted radiomics features, total energy provided the strongest predictive value for both favorable outcome and HT, with AUC of 0.694 (95% CI 0.649 – 0.737] and 0.812 (0.755 – 0.864), respectively. Total energy, energy, maximum, and range emerged as the most influential features in predicting both outcomes, performing comparably with the ischemic lesion volume on NCCT (Table S7, Figure S9). For predicting favorable functional recovery, a logistic regression model incorporating only clinical features worked the best than other models. Its performance was significantly better than the radiomics-based model (AUC of 0.782 vs 0.591, P <0.001). For predicting HT, a random forest model incorporating radiomics features performed the best and significantly outperformed the clinical model (AUC of 0.833 vs 0.626, *P* = 0.003; Table S8, Figure S10).

#### Sensitivity Analysis of the Patients with CT Perfusion

In a subgroup of 269 patients who had both NCCT and CTP at baseline, the Spearman’s ρ between ischemic core volume and early follow-up DWI infarct volume was 0.59 (95% CI, 0.49–0.67) for NCCT and 0.66 (95% CI, 0.58–0.73) for CTP, with no significant difference between them (*P* = 0.23; Figure S11). Regarding functional recovery, however, only 23.8% (5 out of 21) of patients with NCCT-derived core volumes >50 mL achieved favorable outcomes, compared to 42.0% (21 out of 50) of those with CTP-derived cores >50 mL (Figure S12).

## Discussion

In this study, the authors 1) developed and extensively validated an automated model for detecting ischemic lesions on NCCT and 2) evaluated its clinical feasibility and the role of radiomics features on separate LVO patients who had complete recanalization through EVT. The automated model achieved 75.3% sensitivity and 79.1% specificity for ischemic lesion detection in an external validation set. In the feasibility cohort, NCCT lesion volumes showed a strong correlation with early follow-up DWI volumes and were inversely associated with favorable outcomes, outperforming ischemic core volume derived from CTP. Notably, radiomics-based models substantially improved the prediction of HT compared to clinical variables alone.

Although the NCCT is the most prevalent imaging modality for acute stroke care, its limited tissue contrast has historically constrained its sensitivity for early ischemic changes on visual evaluation.^13,14^ Our findings indicate that NCCT nevertheless harbors valuable prognostic information. Specifically, when lesions exceeding 50 mL on NCCT, only 17% of patients achieved good recovery after successful recanalization, whereas 36% of those with large infarcts on CTP. This observation suggests that NCCT may capture more definitive and irreversible tissue damage. Given the impracticality of manually quantifying lesion volume, an automated tool that rapidly quantifies ischemic lesion on NCCT could be highly advantageous.

Radiomics features extracted from NCCT provide additional microstructural insights that may not be fully captured by conventional clinical assessments. In our study, radiomics-based models outperformed clinical-variable models in predicting HT, albeit they were less effective in prognosticating favorable functional recoveries. This discrepancy highlights that radiomics features capture distinct information from the ischemic lesions especially those predisposing to HT. These findings align with previous studies showing that NCCT-based radiomics outperforms clinical indicators alone for predicting HT.^15,16^

It is well established that DWI can detect smaller or earlier stage infarct more reliably than NCCT.^17,18^ In our study, NCCT underestimated infarct volume, especially for small or multifocal lesions, although the correlation between NCCT lesion volume and DWI volume improved with increasing lesion size and time from onset. Notably, NCCT’s correlation with DWI infarct volume exceeded that of CTP after two hours from symptom onset, but it was lower during the first two hours, consistent with the dynamic and evolving nature of early ischemic changes. Sensitivity was also markedly higher in single-lesion cases than in those with multiple scattered lesions, reflecting the inherent challenges of visually detecting subtle or scattered ischemic lesions on NCCT.

The methodological rigor of our study represents a key strength. Ground-truth lesion masks were generated from a large, multicenter dataset of DWI-matched NCCT scans, and five experts independently annotated these images while referencing concomitant DWI and apparent diffusion coefficient sequences. By confining the interval between NCCT and DWI to a narrow window, we minimized ischemic lesion evolution and further improved annotation reliability. The use of consensus masks and volumetric metrics mitigated interobserver variability, facilitating a robust framework for model development and validation.^19^ Furthermore, our findings substantiate prior reports of “ghost cores” on CTP, whereas NCCT more consistently reflected consolidated infarction.^11,20^ Our results imply that NCCT can offer reliable tissue-level insights, which, if enhanced through automated lesion detection and combined with vascular imaging, may enable more accurate individualized prognostication and treatment decision-making.

Our results should be interpreted with caution, and few limitations need to be kept in mind. Although validated in multiple cohorts, the study primarily included Korean patients, potentially limiting generalizability. Due to the limitations of NCCT, predictions for very small lesions may be imperfect, as reflected in the imperfect correlation among experts. Further, the clinical feasibility cohort was restricted to those with complete recanalization, which may not reflect outcomes in patients with partial or failed recanalization. Lastly, DWI volumes beyond the hyperacute window can be influenced by evolving infarct dynamics.

In conclusion, this study presents a robust automated NCCT ischemic lesion detection model from a large, multicenter, expert-annotated dataset. Our findings illustrate that NCCT-based lesion volumes and radiomics features carry substantial prognostic weight for functional recoveries and HT. By providing rapid, quantitative, and clinically relevant information, the automated detection software has the potential to improve acute stroke diagnosis and treatment decision-making, particularly in resource-limited environments.^21^

## Supporting information

STARD checklist

## Supplemental Material

Supplemental Methods

Tables S1–S8

Figure S1-S14

References 1-10

