## Supplementary Material for "Automated Ischemic Stroke Lesion Detection On Non-Contrast Brain CT: A Large-Scale Clinical Feasibility Test"

**List of items**

**Supplemental Methods**

**Figure S1.** Volumetric correlation between lesion volumes predicted by the developed model and experts’ annotations on (A) all cases and (B) cases with lesions.

**Figure S2**. Scatter plots with log scale of the patients receiving non-contrast CT within 180 minutes of symptom onset. (A) Estimated ischemic core non-contrast CT and early follow-up diffusion weighted imaging infarct volume, (B) ischemic core on CT perfusion and early follow-up diffusion weighted imaging infarct volume, and (C) estimated ischemic core non-contrast CT and ischemic core on CT perfusion

**Figure S3**. Scatter plots with log scale of patients receiving non-contrast CT after 180 minutes of symptom onset. (A) Estimated ischemic core non-contrast CT and early follow-up diffusion weighted imaging infarct volume, (B) ischemic core on CT perfusion and early follow-up diffusion weighted imaging infarct volume, and (C) estimated ischemic core non-contrast CT and ischemic core on CT perfusion

**Figure S4.** Scatter plots with log scale of (A) estimated ischemic core non-contrast CT and late follow-up diffusion weighted imaging infarct volume, (B) ischemic core on CT perfusion and late follow-up diffusion weighted imaging infarct volume.

**Figure S5**. 3-month modified Rankin Scale score stratified by estimated ischemic core on non-contrast CT, 0 – <5, 5 – <10, 10 – <20, 20 – <30, 30 – <40, 40 – <50, and ≥ 50mL

**Figure S6**. Restricted cubic spline analysis.

**Figure S7**. Any hemorrhagic transformation on follow-up images stratified by ischemic core volume on non-contrast CT.

**Figure S8.** Histograms of (A) energy, (B) maximum, (C) minimum, (D) median, (E) range, and (F) root mean squared values extracted from the ischemic lesions.

**Figure S9.** Area under the receiver operating characteristic curves and confidence intervals of the radiomic features and non-contrast CT ischemic lesion volume for prediction of (A) favorable outcome and (B) hemorrhagic transformation.

**Figure S10.** Receiver operating characteristic curves on external validation of the machine learning models for prediction of hemorrhagic transformation (A, B) and favorable outcome (C, D) using radiomic features (A, C) and clinical variables (B, D).

**Figure S11.** Scatter plots between (A) estimated ischemic core non-contrast CT and early follow-up diffusion weighted imaging infarct volume, (B) ischemic core on CT perfusion and early follow-up diffusion weighted imaging infarct volume in patients who simultaneously underwent non-contrast CT and CT perfusion.

**Figure S12.** 3-month modified Rankin Scale score stratified by estimated ischemic core on (A) non-contrast CT or (B) CT perfusion, ≥ 50 mL versus < 50 mL in patients who simultaneously underwent non-contrast CT and CT perfusion.

**Figure S13*.*** Structure and output information of 3D-UNet implemented in the study.

**Figure S14.** Inference process of the ischemic lesion detection model.

**Table S1.** Baseline characteristics of dataset for model development

**Table S2.** Volumetric correlation between experts on external validation dataset.

**Table S3.** Performance of the algorithm on external validation dataset stratified based on lesion size

**Table S4.** Performance of the algorithm on external validation dataset stratified based on number of lesions

**Table S5.** Correlation coefficient of ischemic core based on non-contrast CT and CT perfusion in comparison to early follow-up infarct volume on diffusion weighted imaging.

**Table S6.** Multivariable logistic regression analysis between estimated ischemic core volume on non-contrast CT and binary 3-month outcome

**Table S7.** Area under the receiver operating curves for radiomic features in prediction of favorable outcome and hemorrhagic transformation.

**Table S8**. Comparison of the performance of the models using radiomic features and clinical variables for prediction of hemorrhagic transformation and favorable outcome.

**Supplemental Methods**

**Development and Validation of Acute Ischemic Stroke Lesion Detection Software**

**Study subjects**

For the training and internal validation cohort, we retrospectively collected data on consecutive patients with ischemic stroke (admitted between May 2011 and March 2015) from four Korean comprehensive stroke centers with following inclusion criteria: 1) age ≥18 years; 2) admission within 7 days of symptom onset; 3) DWI scan acquired between -1 hour and +3 hours from NCCT scan. Exclusion criteria were as follows: 1) presence of brain tumors, intracranial hemorrhages, ventriculoperitoneal shunts, or extraventricular drains; 2) poor image quality that precluded reliable analysis; and 3) missing annotations from any of the five expert readers.

For the external validation cohort, we retrospectively collected data on consecutive patients with ischemic stroke (admitted between May 2011 and March 2022) from two additional Korean comprehensive stroke centers using the same criteria. Additionally, as a stroke-negative external dataset for the external validation cohort, consecutive patients admitted between January 2017 and December 2020 from a comprehensive stroke center were included. Patients in the negative control initially presented with stroke-like symptoms and underwent NCCT imaging, but no ischemic lesions were found on subsequent DWI scans. Acute ischemic stroke was subsequently ruled out by consensus among experienced neurologists and radiologists. We retrieved baseline demographic and clinical information for all study participants using a standardized protocol.^1^ The Institutional Review Board approved the study and waived informed consent requirements owing to its retrospective design and de-identified data (IRB approval # B-2102-667-106).

**Ischemic stroke lesion segmentation**

Five experienced neurologists from South Korea, with > 10 years of clinical experience independently delineated the hypoattenuation area corresponding to acute ischemic changes on the baseline NCCT scans by referring to corresponding follow-up DWI and apparent diffusion coefficient (ADC) images. They were instructed to forego annotation if NCCT did not exhibit any definite infarct-related changes, even if ischemic infarcts were evident on DWI. Aside from NCCT and DWI/ADC series, no additional clinical information was provided to the raters. All segmentation tasks were carried out using an in-house platform (Medihub Stroke, JLK, Korea), and all images were subsequently reviewed and validated through inter-rater consensus.

**Data Preparation**

To reduce the impact of irrelevant features such as background elements and gantry tilts from the CT scanner, we cropped out extraneous spaces and applied padding with value of -1000HU to maintain a square dimension. On four distinct models, we employed different Hounsfield Unit windowing settings followed by min-max scaling: [W:80, L:40], [W:40, L:40], [W:60, L:35], and [W:30, L:35], respectively. Non-contrast CT (NCCT) images and corresponding hypodensity masks provided by experts were resized to a resolution of 256x256 pixels using bilinear interpolation. Random rotation with ranges of -30 to 30 degree, horizontal flipping, translation, brightness adjustment, and scaling were applied for data augmentation.

**Model Architecture and Training**

We employed a modified version of 3D U-Net for hypodense lesion segmentation.^2^ While preserving its U-shaped architecture, several parameters, basic convolution blocks, and skip connection methods were modified. The model includes 6 stages with multiple convolutional layers per stage together with down-sampling and up-sampling block for encoder and decoder, respectively. A Leaky ReLU activation function with a negative slope of 0.01 was employed. The training regimen included a hybrid loss function combining Dice loss and binary cross-entropy with equal weighting, a batch size of 8, AdamW optimizer incorporating weight decay, and a cosine learning rate decay with an initial warm-up phase, executed on a single Nvidia RTX A6000 GPU using Pytorch.

To simulate variability of lesion identification, the model was trained to handle multi-label segmentation, producing five independent segmentation outputs based on annotations from five experts. As an input, 3D patches with dimension of 20x256x256 were extracted from a preprocessed NCCT images. For each patch, the model outputs a pixel-wise sigmoid probability map with dimensions 5x20x256x256, where the first dimension represents each expert’s segmentation (Supplementary Figure 12). The final segmentation output is generated by averaging the pixel-wise probabilities across the five expert-specific outputs, consolidating multi-expert insights. The convolution kernel sizes were (1×3×3), (1×3×3), (3×3×3), (3×3×3), (3×3×3), and (3×3×3), while the down sampling kernel sizes were (1,2,2), (1,2,2), (2,2,2), (2,2,2), and (1,2,2).

We enhanced model robustness through an ensemble approach, incorporating models trained on data with four distinct Hounsfield Unit windowing presets. Hyperparameters were standardized across all ensemble models, and a fixed random seed controlled the randomness of data loading and model initialization. For each model, final selection was done based on Dice Similarity Coefficient (DSC) on internal validation dataset.

**Inference Process of the Model**

For inference, multiple patches with 50% overlap on z axis were extracted from NCCT images of a single case. The outputs of input patches, represented as pixel-wise sigmoid probability, are added and divided by number of inferences (Supplementary Figure 13). The final outputs were thresholded at 0.5; pixels greater than or equal to 0.5 are set to 1, while others less than 0.5 are set to 0.

**Evaluation of the Ischemic Stroke Lesion Detection Model**

Before evaluating the model, we assessed the agreement between experts on their annotations using volumetric similarity (VS) and absolute volume difference (AVD). Ground truth (GT) masks for evaluation were defined as voxels agreed upon by more than two experts. Considering the known limitation in detecting extremely small lesions on standard NCCT images, we excluded any lesion with a volume under 0.5 mL in both the ground truth and predicted masks during evaluation. Therefore, in the external validation, any ischemic lesion exceeding 0.5 mL on DWI was designated as positive. DSC was only calculated on cases where GT masks include any lesion to avoid misinterpretation. True positives only included cases that had any overlap between prediction and GT masks. Also, we assessed the model’s performance depending on the volume and number of GT lesions on NCCT.

**Clinical Application of the Automated Ischemic Stroke Lesion Detection Software**

**Clinical Data Collection**

Included baseline demographic and clinical information were age, sex, history of previous stroke, functional status before stroke, and cardiovascular risk factors such as hypertension, diabetes mellitus, and atrial fibrillation. Stroke characteristics included the time interval between last known well and NCCT scan, the National Institutes of Health Stroke Scale (NIHSS) score at admission, and treatment information. Admission NIHSS scores, 3-month modified Rankin Scale (mRS) scores were evaluated by certified (http://www.stroke-edu.or.kr/) physicians or stroke coordinators in each stroke center.^3^

**Radiomic features extracted from the ischemic stroke lesions**

Energy and total energy represent the overall intensity of voxel values within the region of interest, reflecting signal strength. Entropy quantifies randomness in the intensity distribution, indicating texture complexity. Minimum, 10th percentile, 90th percentile, and maximum capture the range of intensity values, highlighting extremes and distribution spread. Mean and median represent central tendencies, with median being less sensitive to outliers. Interquartile range and range describe variability, with the former focusing on the middle 50% of values. Mean absolute deviation and robust mean absolute deviation measure dispersion, with the latter being less influenced by outliers. Root mean squared assesses the magnitude of voxel values, incorporating both variance and mean intensity. Skewness indicates asymmetry in the intensity distribution, while kurtosis describes the presence of extreme values. Variance quantifies overall dispersion, and uniformity measures the consistency of intensity values, with higher values indicating more homogeneity. The features were standardized to normalize numerical values before model training. No missing values were present in the dataset.

**Outcome Prediction using Machine Learning with Radiomic Features**

Machine learning models to predict favorable outcomes and hemorrhagic transformation were developed and validated with radiomic features extracted from the automated ischemic lesion extracted from the software. Patients who had any lesion in NCCT were included from the two cohorts of the clinical application dataset. Patients in the first cohort were included for model development, and patients in the second cohort were included as the external validation dataset. From these patients, a total of 18 first-order radiomics features were extracted with the *PyRadiomics* package.^4^ The description of the radiomics features is presented in the Supplementary Methods. Hemorrhagic transformation and 3-month mRS score were dichotomized as outcomes for the prediction models. Favorable outcome was defined as 3-month mRS ≤2.^5^ Hemorrhagic transformation was dichotomized as no hemorrhagic transformation or hemorrhagic infarction (type 1 or 2) vs. parenchymal hemorrhage (type 1 or 2). The predictive performance of each radiomics feature to predict favorable outcome and hemorrhagic transformation was assessed with the area-under-the-curve of receiver operating characteristics.

**Training and Validation of the Radiomics Machine Learning Model**

To evaluate the predictive performance of the radiomic features compared to clinical variables on outcomes, separate machine learning models were developed using clinical variables and radiomics features. The models were trained with the development dataset, randomly split into training and internal validation in an 8:2 ratio. Five machine learning algorithms were implemented: logistic regression, random forest, XGBoost, LightGBM, and Gradient Boosting Machine. Hyperparameter tuning was performed for each model using a grid search with 5-fold cross-validation, optimizing for area under the receiver operating characteristic curve (AUC) scores. Using the model with the best-performing hyperparameters, AUC scores were calculated for the training, internal validation, and external validation datasets. The models were trained with Python (version 3.9.18, Python Software Foundation, Delaware, USA) using *Scikit-learn*, *XGBoost*, and *LightGBM* packages.^6–8^

**Statistical analysis**

Data were presented as mean±standard deviation, median (interquartile range), or number (percentage). For comparison of baseline characteristics between training, internal and external validation datasets, we used ANOVA or Kruskal-Wallis test for continuous variables and chi-square test or Fisher exact test for categorical variables as appropriate.

To compare the ischemic core volume estimated by the algorithm on NCCT with the ischemic core on CT perfusion and with early and late ischemic lesions on diffusion weighted imaging, we calculated Spearman’s correlation considering the skewed distribution of infarct volume. Correlation coefficients were compared using Fisher’s z-transformation.^9^ The analysis was also repeated after stratifying by onset-to-NCCT scan time (≤3 hours vs. >3 hours). Based on predefined thresholds, the ischemic core volume on NCCT estimated by the algorithm or on CT perfusion was categorized as <50 mL vs. ≥50 mL. Further, for a more granular classification, the ischemic core volume on NCCT was categorized at 0, 5, 10, 20, 30, 40, and 50 mL increments.

Associations between these categorized ischemic core volumes on NCCT, 3-month mRS scores, and European Cooperative Acute Stroke Study II hemorrhagic transformation were evaluated using the chi-square test. Subsequently, a multivariable binary logistic regression was conducted to assess the independent association of categorized ischemic core volume on NCCT with favorable outcome (3-month mRS score ≤2). Covariates were chosen based on prior research and included age, sex, onset-to-NCCT scan time, NCCT-to-groin puncture time, endovascular thrombectomy procedure time, intravenous thrombolysis, previous stroke, coronary artery disease, hypertension, diabetes, hyperlipidemia, smoking, and atrial fibrillation.^10^ A restricted cubic spline was employed to explore possible non-linear relationships between NCCT ischemic core volume and favorable 3-month outcomes.

In addition to the AUCs of the machine learning models for prediction of outcomes, PPV, NPV, Akaike information criterion, Bayesian information criterion, sensitivity, and specificity were analyzed. Youden’s index was used for determining the optimal cut-offs. DeLong’s test was used for comparing AUCs between the models.

For sensitivity analyses, we restricted the sample to patients who underwent both NCCT and CT perfusion simultaneously, comparing NCCT- and CT perfusion-derived ischemic core volumes with early follow-up infarct volume and evaluating the relationship between an NCCT ischemic core threshold of 50 mL and 3-month outcomes.

A two-tailed p-value <0.05 was considered statistically significant. Statistical analyses were performed using STATA (version 16.0, StataCorp, College Station, TX), MedCalc Statistical Software (version 22.016, MedCalc Software, Mariakerke, Belgium), R (version 4.3.3, R Foundation for Statistical Computing, Vienna, Austria), and Python (version 3.9.18, Python Software Foundation, Delaware, USA).

**Figure S1. Volumetric correlation between lesion volumes predicted by the developed model and experts’ annotations on (A) all cases and (B) cases with lesions.**

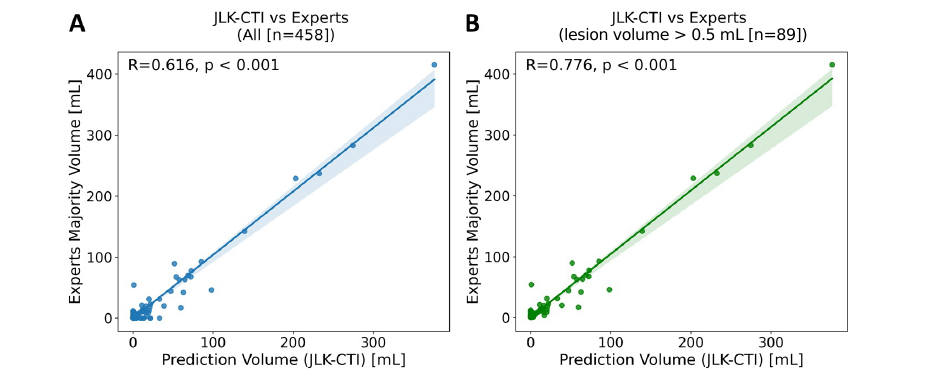

**Figure S2. Scatter plots with log scale of the patients receiving non-contrast CT within 180 minutes of symptom onset. (A) Estimated ischemic core non-contrast CT and early follow-up diffusion weighted imaging infarct volume, (B) ischemic core on CT perfusion and early follow-up diffusion weighted imaging infarct volume, and (C) estimated ischemic core non-contrast CT and ischemic core on CT perfusion**

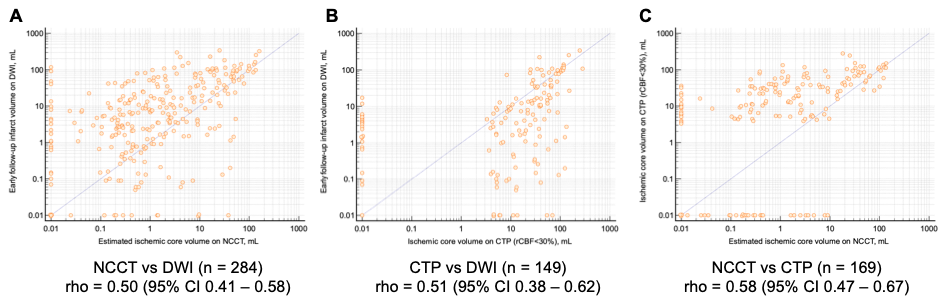

NCCT, non-contrast CT; DWI, diffusion-weighted imaging; CTP, CT perfusion; CI, confidence interval.

**Figure S3. Scatter plots with log scale of patients receiving non-contrast CT after 180 minutes of symptom onset. (A) Estimated ischemic core non-contrast CT and early follow-up diffusion weighted imaging infarct volume, (B) ischemic core on CT perfusion and early follow-up diffusion weighted imaging infarct volume, and (C) estimated ischemic core non-contrast CT and ischemic core on CT perfusion**

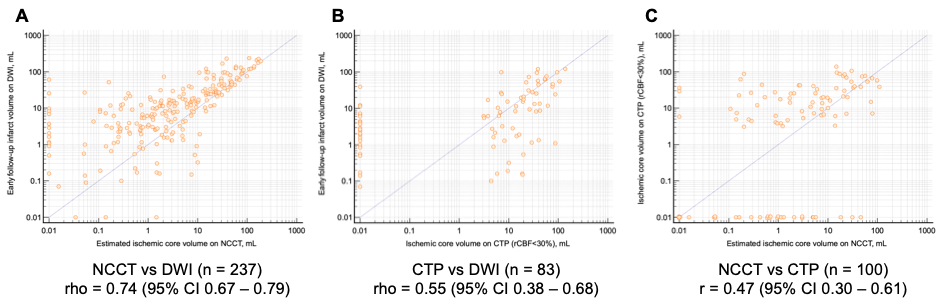

NCCT, non-contrast CT; DWI, diffusion-weighted imaging; CTP, CT perfusion; CI, confidence interval.

**Figure S4. Scatter plots with log scale of (A) estimated ischemic core non-contrast CT and late follow-up diffusion weighted imaging infarct volume, (B) ischemic core on CT perfusion and late follow-up diffusion weighted imaging infarct volume.**

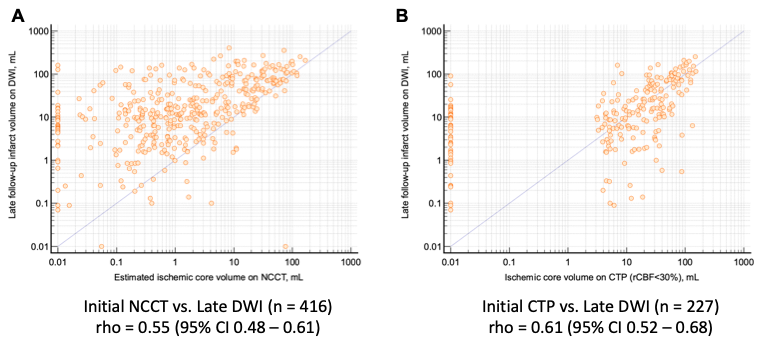

NCCT, non-contrast CT; DWI, diffusion-weighted imaging; CTP, CT perfusion; CI, confidence interval.

**Figure S5. 3-month modified Rankin Scale score stratified by estimated ischemic core on non-contrast CT, 0 – <5, 5 – <10, 10 – <20, 20 – <30, 30 – <40, 40 – <50, and ≥ 50mL**

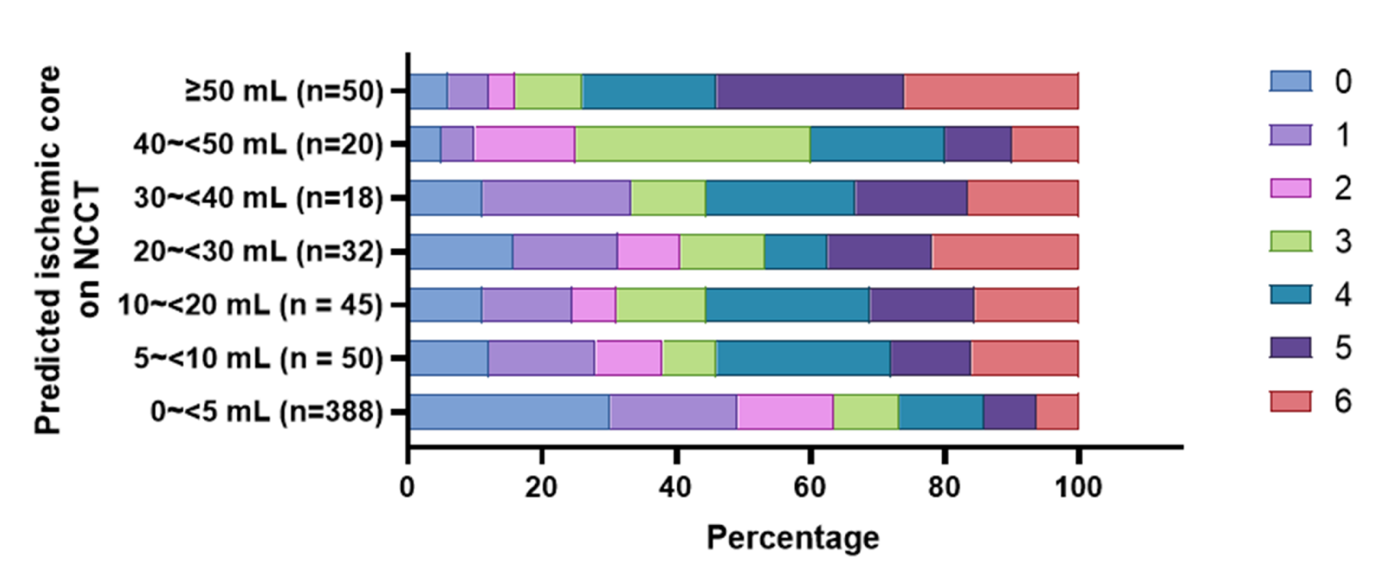

NCCT, non-contrast CT

**Figure S6. Restricted cubic spline analysis.**

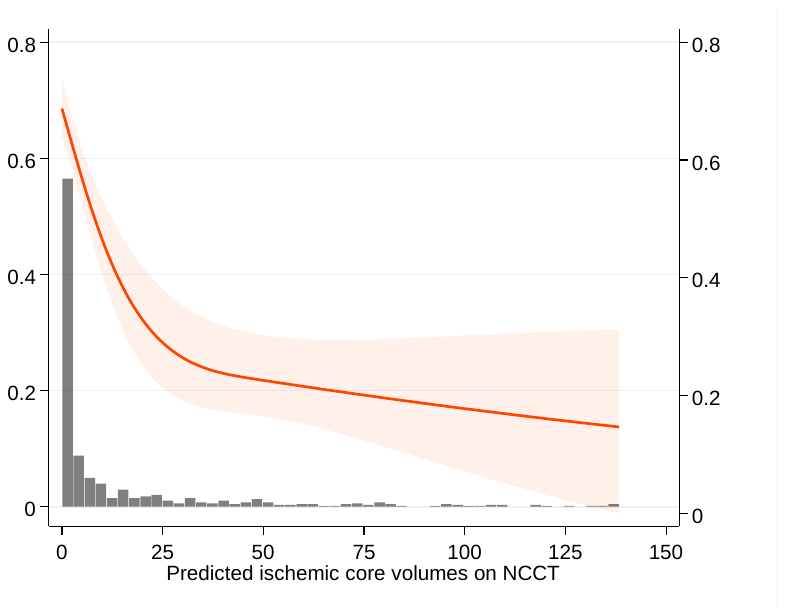

NCCT, non-contrast CT

**Figure S7. Any hemorrhagic transformation on follow-up images stratified by ischemic core volume on non-contrast CT.**

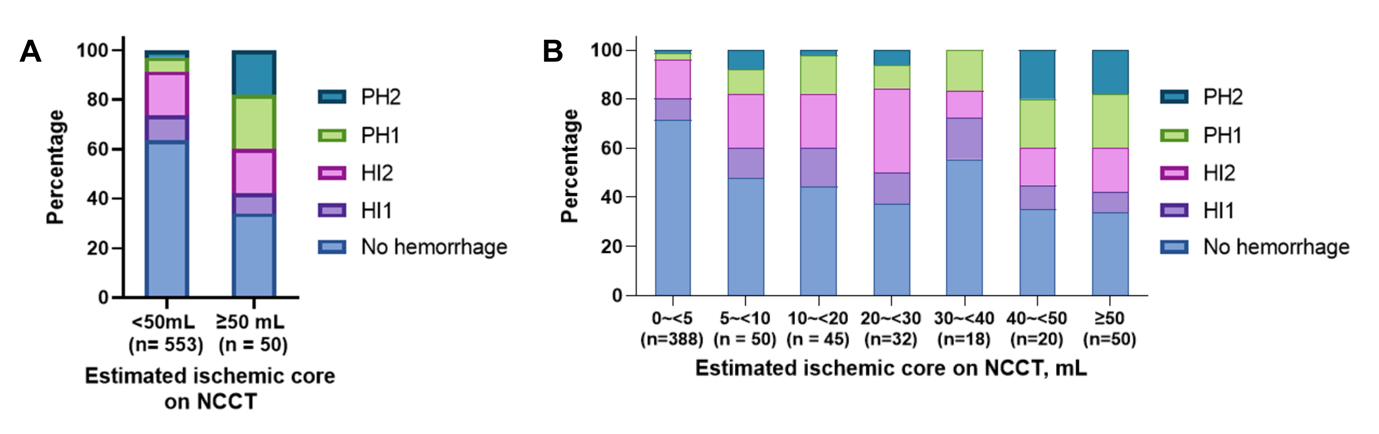

HI, hemorrhagic infarct; PH, parenchymal hemorrhage; NCCT, non-contrast CT.

**Figure S8. Histograms of (A) energy, (B) maximum, (C) minimum, (D) median, (E) range, and (F) root mean squared values extracted from the ischemic lesions.**
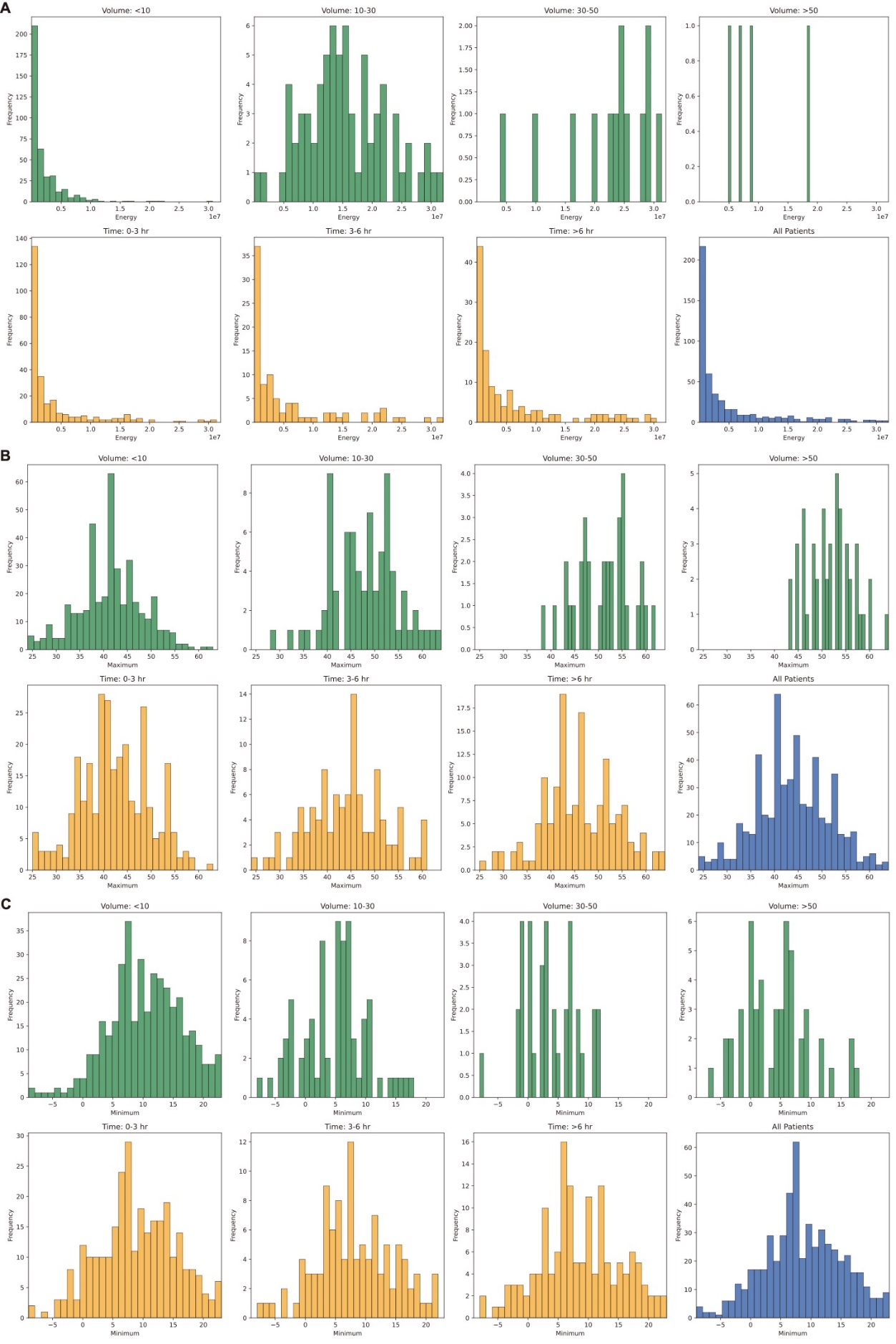

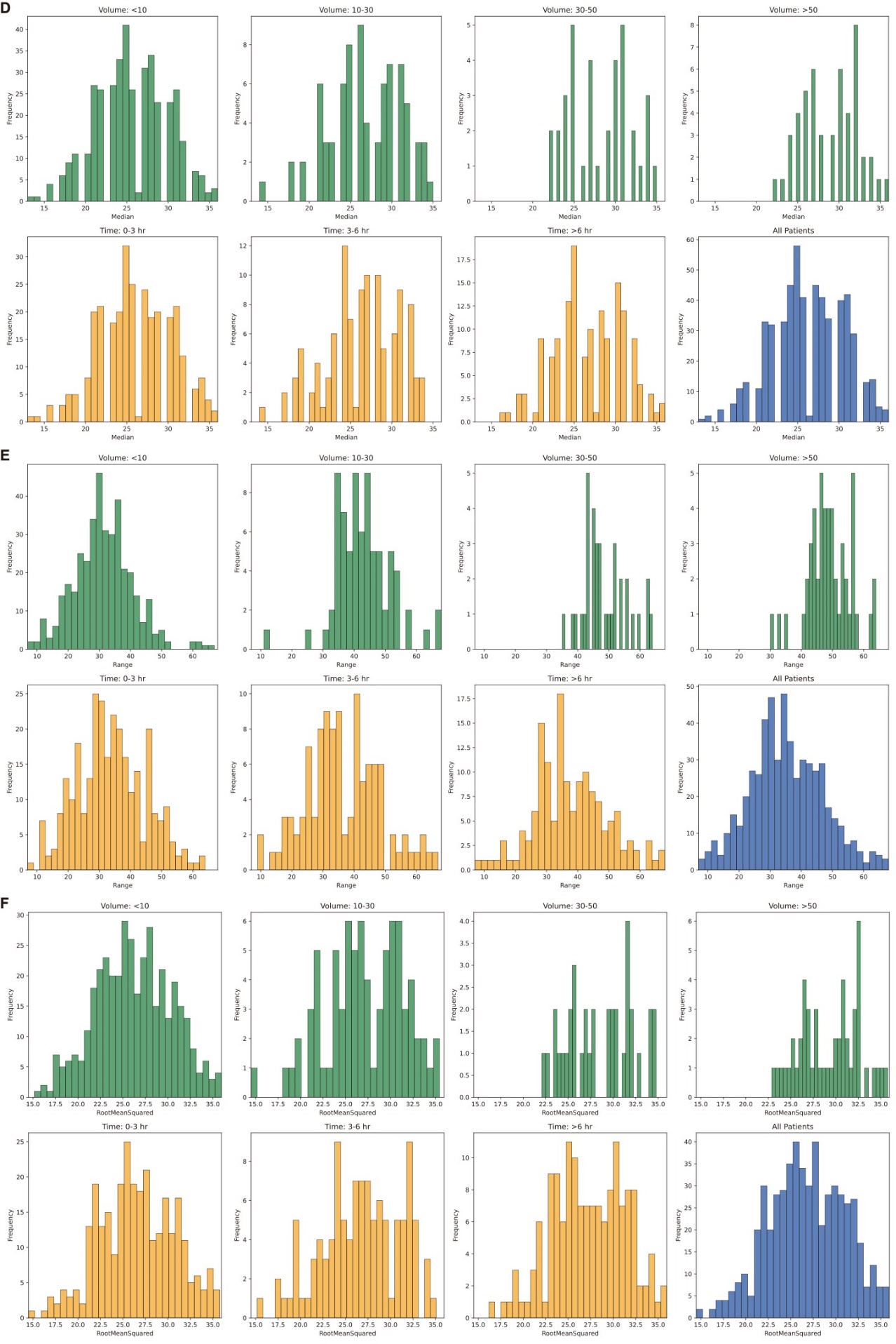

Green colors were used for graphs showing volume subgroups, yellow for time subgroups, and blue for graphs including all patients.

**Figure S9. Area under the receiver operating characteristic curves and confidence intervals of the radiomic features and non-contrast CT ischemic lesion volume for prediction of (A) favorable outcome and (B) hemorrhagic transformation.**

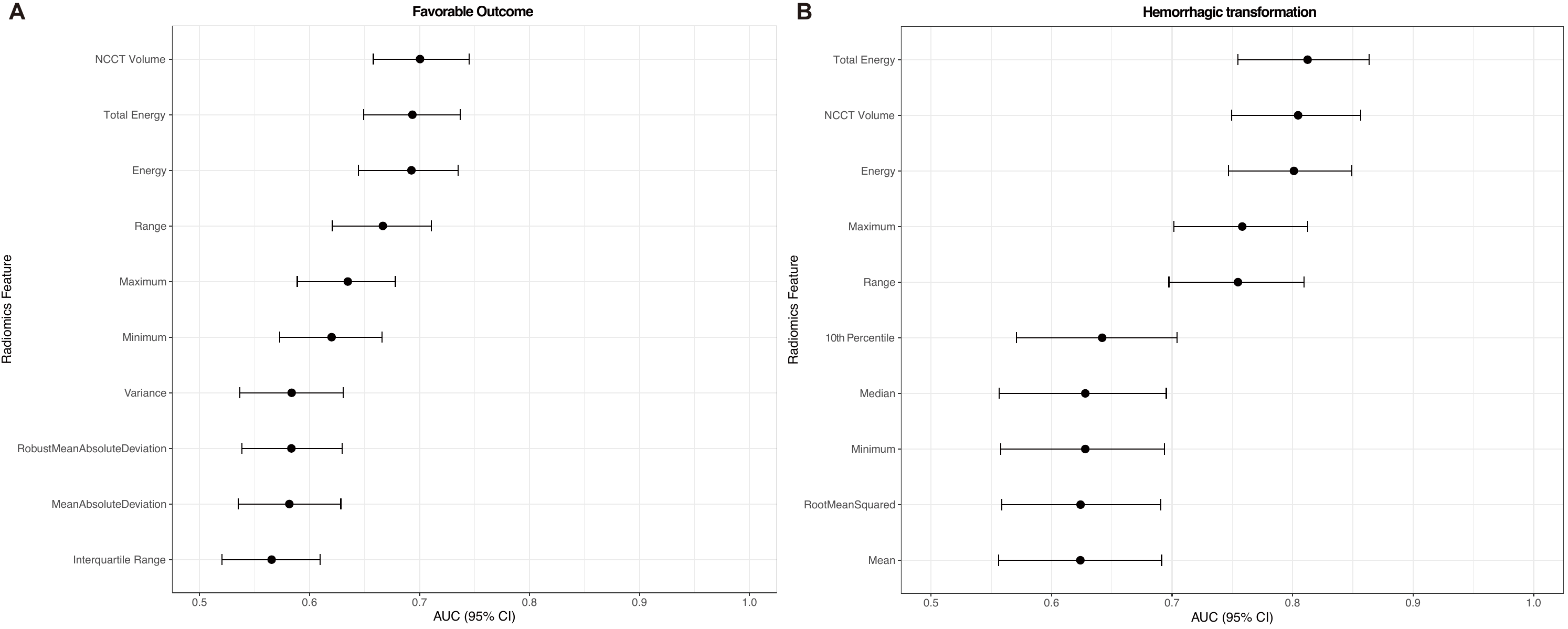

NCCT, non-contrast CT; AUC, area under the curve receiver operating characteristics; CI, confidence interval.

**Figure S10.** **Receiver operating characteristic curves on external validation of the machine learning models for prediction of hemorrhagic transformation (A, B) and favorable outcome (C, D) using radiomic features (A, C) and clinical variables (B, D).**

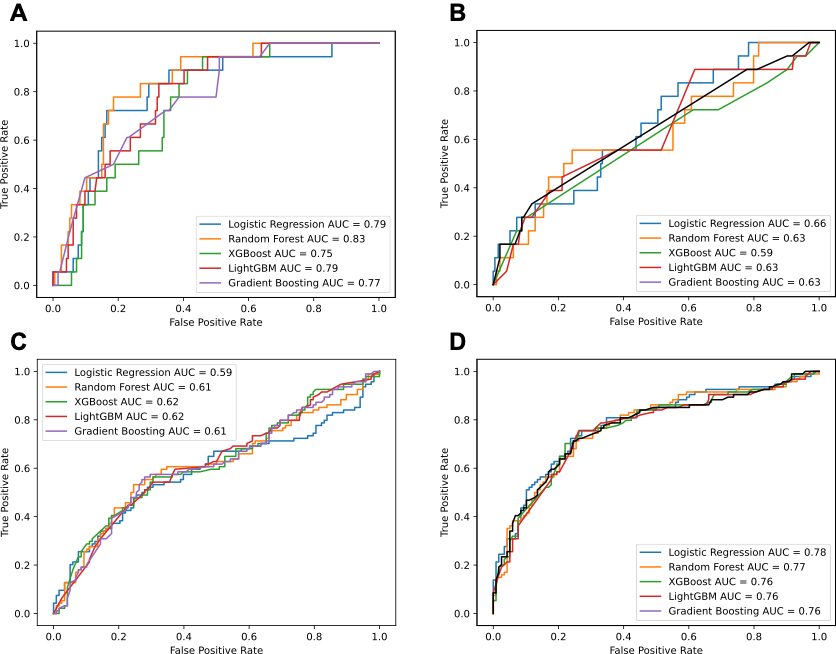

AUC, area under the curve receiver operating characteristics

**Figure S11. Scatter plots between (A) estimated ischemic core non-contrast CT and early follow-up diffusion weighted imaging infarct volume, (B) ischemic core on CT perfusion and early follow-up diffusion weighted imaging infarct volume in patients who simultaneously underwent non-contrast CT and CT perfusion.**

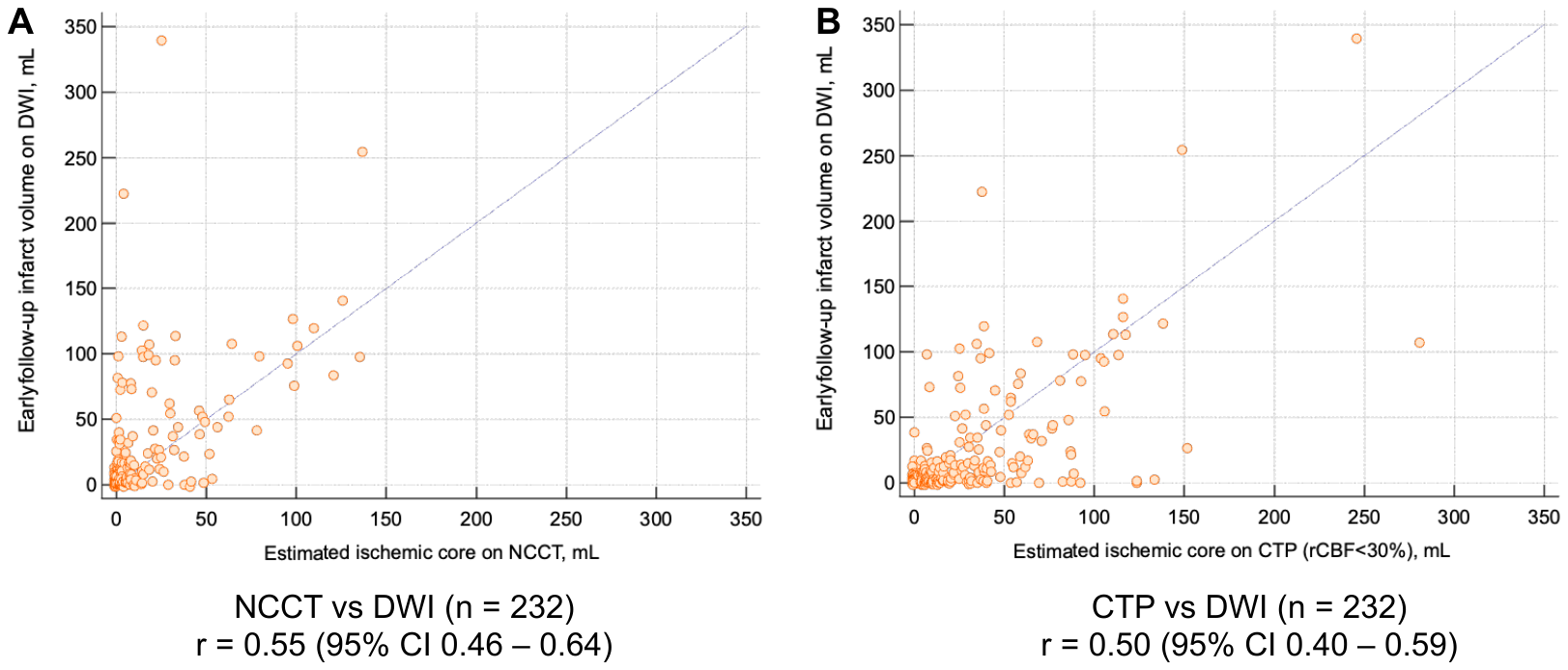

NCCT, non-contrast CT; DWI, diffusion-weighted imaging; CTP, CT perfusion; CI, confidence interval.

**Figure S12. 3-month modified Rankin Scale score stratified by estimated ischemic core on (A) non-contrast CT or (B) CT perfusion, ≥ 50 mL versus < 50 mL in patients who simultaneously underwent non-contrast CT and CT perfusion.**

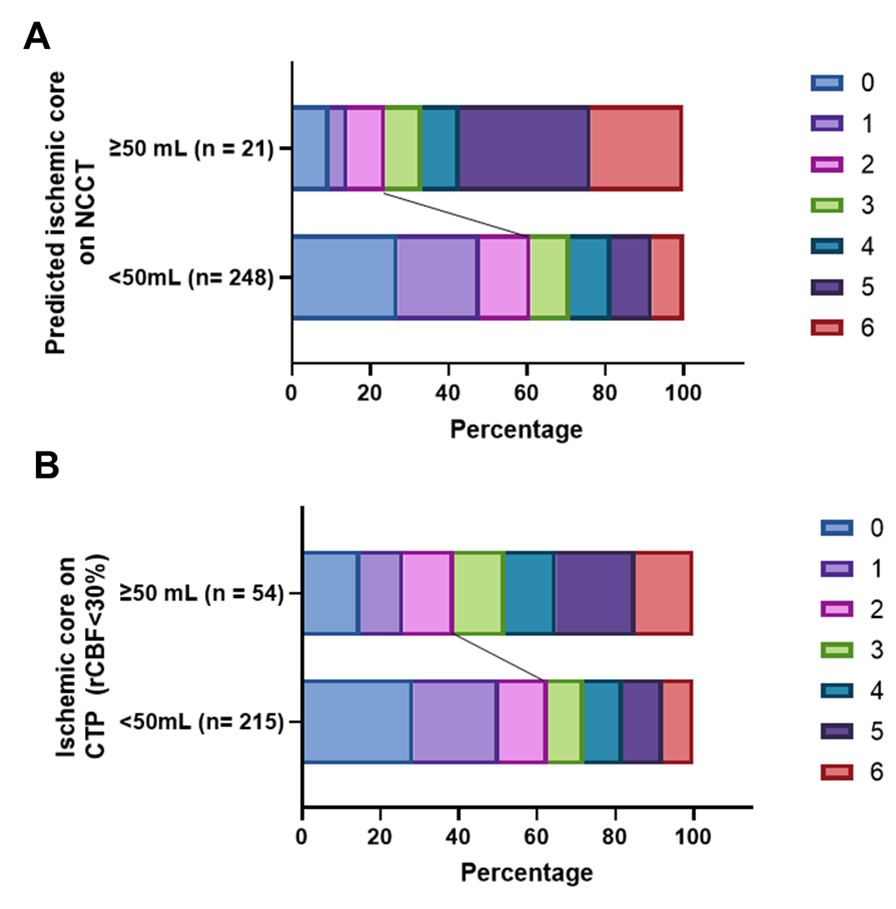

NCCT, non-contrast CT; CTP, CT perfusion; rCBF, regional cerebral blood flow.

**Figure S13*.* Structure and output information of 3D-UNet implemented in the study.**

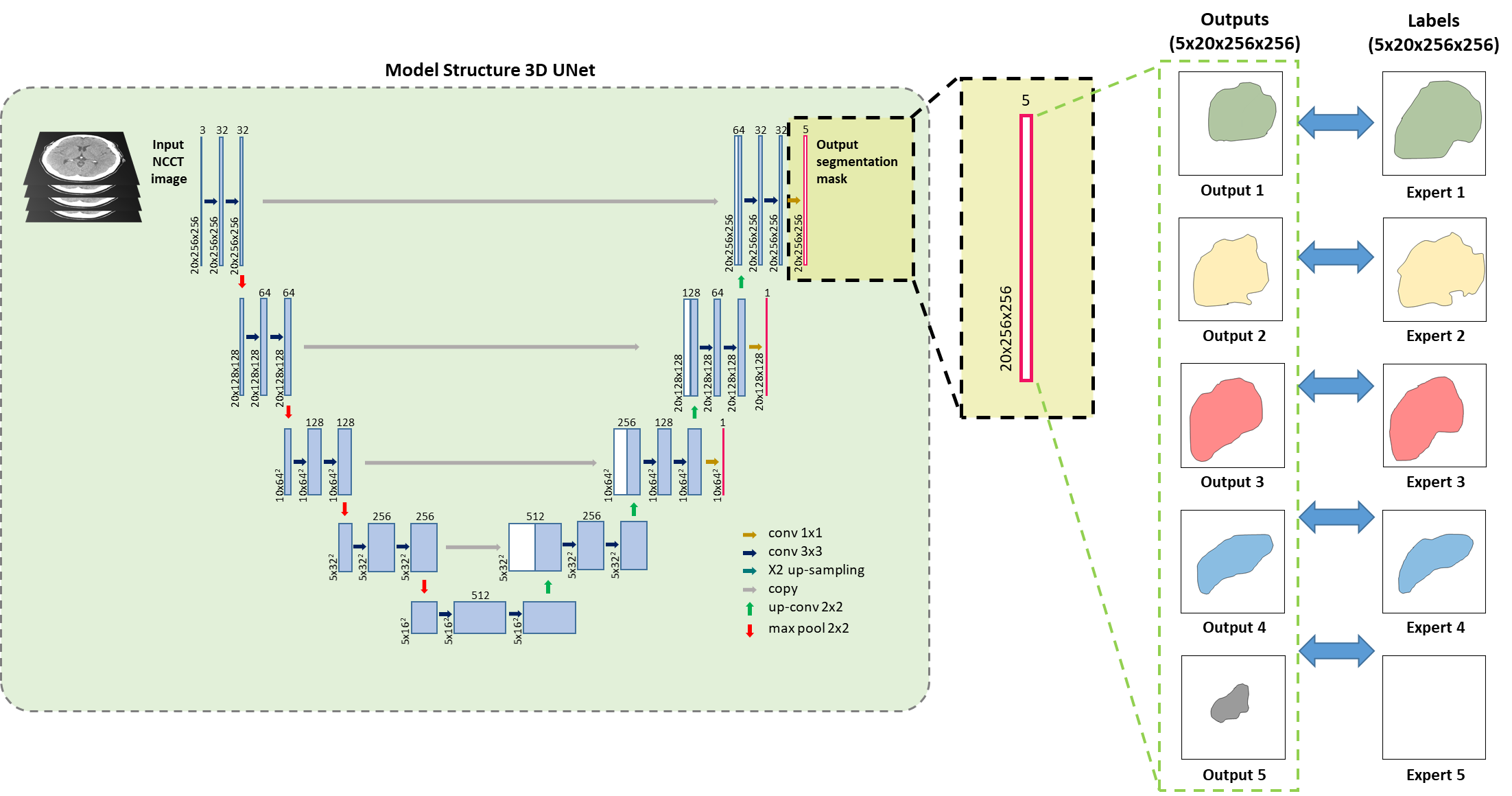

NCCT, non-contrast CT

**Figure S14. Inference process of the ischemic lesion detection model.**

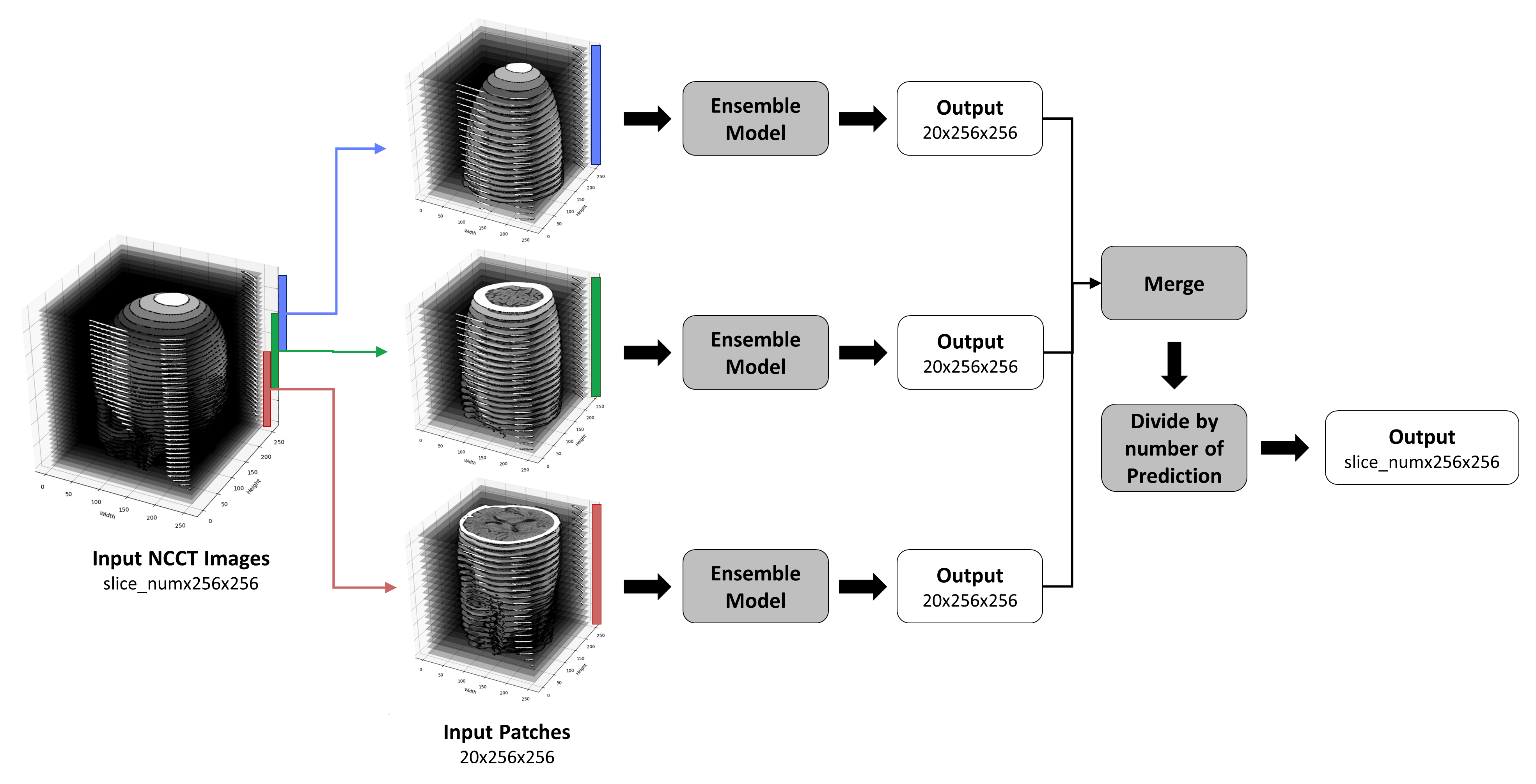

NCCT, non-contrast CT

**Table S1. Baseline characteristics of datasets for model development**

|  | Model development and training set  (n = 1,991) | Internal validation set  (n = 223) | External validation set  (n = 458) | P value |
| --- | --- | --- | --- | --- |
| Age | 69.0±12.2 | 68.5±12.1 | 64.1±15.5 | < 0.001 |
| Male | 1,136 (57.3%) | 144 (64.6%) | 275 (60.3%) | 0.079 |
| Time between NCCT and DWI, hrs | 0.50 (0.29–0.87) | 0.43 (0.28–0.83) | 0.56 (0.35–0.84) | 0.17 |
| Vendor |  |  |  | < 0.001 |
| GE | 687 (34.5%) | 80 (35.9%) | 0 |  |
| Philips | 21 (1.1%) | 1 (0.5%) | 232 (50.7%) |  |
| SIEMENS | 1,247 (62.6%) | 138 (61.9%) | 226 (49.3%) |  |
| Kvp |  |  |  | < 0.001 |
| 100 | 156 (7.8%) | 17 (7.6%) | 44 (9.6%) |  |
| 120 | 1,819 (91.4%) | 205 (91.9%) | 414 (90.4%) |  |
| Slice thickness |  |  |  | < 0.001 |
| ≤ 4 mm | 436 (21.9%) | 50 (22.4%) | 1 (0.2%) |  |
| 4.8 mm | 811 (40.7%) | 90 (40.4%) | 108 (23.6%) |  |
| 5 mm | 744 (37.4%) | 83 (37.2%) | 349 (76.0%) |  |
| Onset to image, hr | 8.5 (2.7 – 20.5) | 8.7 (3.2 – 24.7) | 8.8 (2 – 25.3)* | 0.59 |

*Data were available for 184 patients with ischemic stroke.

NCCT. noncontrast CT; DWI, diffusion-weighted imaging

**Table S2. Volumetric correlation between experts on external validation dataset.**

|  | Expert 1 | Expert 2 | Expert 3 | Expert 4 | Expert 5 |
| --- | --- | --- | --- | --- | --- |
| Expert 1 | 1 | 0.766 | 0.807 | 0.753 | 0.748 |
| Expert 2 |  | 1 | 0.828 | 0.868 | 0.841 |
| Expert 3 |  |  | 1 | 0.861 | 0.841 |
| Expert 4 |  |  |  | 1 | 0.873 |
| Expert 5 |  |  |  |  | 1 |

**Table S3. Performance of the algorithm on external validation dataset stratified based on lesion size**

| **metrics** | **All**  (n=89) | **0.5 – 1 mL**  (n=15) | **1 - <5 mL**  (n=27) | **5 - <10 mL**  (n=11) | **10 - <30 mL**  (n=17) | **>30 mL**  (n=19) |
| --- | --- | --- | --- | --- | --- | --- |
| Sens | 0.753 (0.709 - 0.799) | 0.533 (0.412 - 0.667) | 0.667 (0.582 - 0.762) | 0.727 (0.596 - 0.860) | 0.882 (0.807 - 0.961) | 0.947 (0.897 - 0.999) |
| DSC | 0.509 (0.470 - 0.546) | 0.357 (0.264 - 0.456) | 0.351 (0.289 - 0.411) | 0.429 (0.331 - 0.528) | 0.651 (0.58 - 0.728) | 0.775 (0.725 - 0.827) |

Sens, sensitivity; DSC, Dice similarity coefficient

**Table S4. Performance of the algorithm on external validation dataset stratified based on the count of lesions**

| **metrics** | **Lesion counts** | **All**  (n=37, single lesion)  (n=52, multiple lesions) | $\boldsymbol{\leq}$**5 mL**  (n=19, single lesion)  (n=23, multiple lesions) | **5 – 30 mL**  (n=9, single lesion)  (n=19, multiple lesions) | **>30 mL**  (n=9, single lesion)  (n=10, multiple lesions) |
| --- | --- | --- | --- | --- | --- |
| Sens | Single | 0.919 (0.871 - 0.963) | 0.842 (0.761 - 0.923) | 1.000 (1.000 - 1.000) | 1.000 (1.000 - 1.000) |
|  | Multiple | 0.635 (0.566 - 0.699) | 0.435 (0.328 - 0.538) | 0.737 (0.638 - 0.843) | 0.900 (0.805 - 0.996) |
| DSC | Single | 0.687 (0.641 - 0.731) | 0.571 (0.501 - 0.636) | 0.786 (0.741 - 0.835) | 0.832 (0.789 - 0.874) |
|  | Multiple | 0.383 (0.334 - 0.432) | 0.173 (0.121 - 0.223) | 0.458 (0.381 - 0.540) | 0.724 (0.635 - 0.808) |

Sens, sensitivity; DSC, Dice similarity coefficient

**Table S5. Correlation coefficient of ischemic core based on non-contrast CT and CT perfusion in comparison to early follow-up infarct volume on diffusion-weighted imaging.**

|  |  | All | >1 hour | >2 hours | >3 hours | >4 hours | >5 hours | >6 hours |
| --- | --- | --- | --- | --- | --- | --- | --- | --- |
| Number of patients | NCCT | 520 | 442 | 303 | 237 | 190 | 154 | 141 |
|  | CTP | 231 | 186 | 119 | 83 | 61 | 49 | 44 |
| Correlation (95% CI) | NCCT | 0.68 (0.63 to 0.72) | 0.69 (0.64 to 0.74) | 0.73 (0.67 to 0.78) | 0.84 (0.80 to 0.87) | 0.84 (0.80 to 0.88) | 0.85 (0.79 to 0.89) | 0.85 (0.80 to 0.89) |
|  | CTP | 0.64 (0.56 to 0.71) | 0.67 (0.59 to 0.75) | 0.57 (0.43 to 0.68) | 0.61 (0.46 to 0.73) | 0.57 (0.37 to 0.72) | 0.56 (0.34 to 0.73) | 0.53 (0.28 to 0.72) |
|  | P for difference | 0.37 | 0.67 | 0.01 | < 0.001 | < 0.001 | < 0.001 | < 0.001 |

NCCT, non-contrast CT; CTP, CT perfusion; CI, confidence intervals.

**Table S6. Multivariable logistic regression analysis between estimated ischemic core volume on non-contrast CT and binary 3-month outcome**

|  | Unadjusted odds ratio (95% CI) | Adjusted^a^ odds ratio (95% CI) |
| --- | --- | --- |
| Continuous, per 1 mL increase | 1.03 (1.02–1.04) | 1.03 (1.02–1.04) |
| Category |  |  |
| 0 - <5mL (n = 391) | Reference | Reference |
| 5 – <10 mL (n = 50) | 2.77 (1.51 – 5.07) | 2.70 (1.36 – 5.36) |
| 10 – <20 mL (n = 45) | 3.76 (1.93 – 7.29) | 5.63 (2.67 – 11.90) |
| 20 – <30 mL (n = 34) | 2.74 (1.33 – 5.64) | 2.68 (1.16 – 6.20) |
| 30 – <40 mL (n = 18) | 3.39 (1.25 – 9.23) | 4.89 (1.61 – 14.85) |
| 40 – <50 mL (n = 21) | 5.42 (1.94 – 15.13) | 9.60 (3.04 – 30.33) |
| ≥ 50 mL (n = 52) | 8.11 (3.84 – 17.11) | 13.81 (5.78 – 32.98) |

^a^Adjusted for age, sex, onset to NCCT, NCCT to groin puncture, procedure time, intravenous thrombolysis, previous stroke, coronary artery disease, hypertension, diabetes, hyperlipidemia, smoking, and atrial fibrillation

**Table S7. Areas under the curves receiver operating characteristics for radiomic features in prediction of favorable outcome and hemorrhagic transformation.**

| Radiomic features | Favorable outcome | Hemorrhagic transformation |
| --- | --- | --- |
| Energy | 0.693 [0.645 - 0.735] | 0.801 [0.747 - 0.849] |
| Entropy | 0.499 [0.451 - 0.546] | 0.519 [0.443 - 0.594] |
| IQR | 0.566 [0.520 - 0.610] | 0.585 [0.520 - 0.645] |
| Kurtosis | 0.536 [0.489 - 0.584] | 0.605 [0.533 - 0.675] |
| Maximum | 0.635 [0.589 - 0.678] | 0.758 [0.702 - 0.813] |
| Mean | 0.519 [0.471 - 0.566] | 0.624 [0.556 - 0.691] |
| MeanAbsoluteDeviation | 0.582 [0.535 - 0.628] | 0.600 [0.540 - 0.661] |
| Median | 0.521 [0.476 - 0.566] | 0.628 [0.556 - 0.695] |
| Minimum | 0.700 [0.658 - 0.745] | 0.805 [0.750 - 0.857] |
| 10^th^ Percentile | 0.511 [0.460 - 0.558] | 0.585 [0.515 - 0.656] |
| 90^th^ Percentile | 0.542 [0.493 - 0.592] | 0.642 [0.571 - 0.704] |
| Range | 0.667 [0.621 - 0.711] | 0.755 [0.697 - 0.809] |
| RobustMeanAbsoluteDeviation | 0.583 [0.539 - 0.630] | 0.596 [0.535 - 0.655] |
| RootMeanSquared | 0.530 [0.479 - 0.577] | 0.624 [0.559 - 0.691] |
| Skewness | 0.535 [0.486 - 0.582] | 0.573 [0.504 - 0.639] |
| Total Energy | 0.694 [0.649 - 0.737] | 0.812 [0.755 - 0.864] |
| Uniformity | 0.501 [0.451 - 0.549] | 0.519 [0.441 - 0.598] |
| Variance | 0.584 [0.537 - 0.630] | 0.604 [0.541 - 0.665] |
| NCCT Volume (mL) | 0.511 [0.460 - 0.558] | 0.585 [0.515 - 0.656] |

Area under the receiver operating curve is presented as value [95% confidence intervals].

IQR = interquartile range; NCCT = non-contrast CT.

**Table S8. Comparison of the performance of the models using radiomic features and clinical variables for prediction of hemorrhagic transformation and favorable outcome.**

| Model | Predictor | AUC | P-value | AIC | BIC | PPV | NPV | Sensitivity | Specificity |
| --- | --- | --- | --- | --- | --- | --- | --- | --- | --- |
| Favorable outcome | | | | | | | | | |
| GBM | Clinical | 0.767 [0.698-0.832] | 0.002 | 265 | 272 | 0.698 | 0.767 | 0.713 | 0.754 |
|  | Radiomics | 0.614 [0.527-0.685] |  | 309 | 316 | 0.616 | 0.675 | 0.564 | 0.720 |
| LightGBM | Clinical | 0.760 [0.684-0.824] | 0.005 | 272 | 279 | 0.696 | 0.791 | 0.755 | 0.737 |
|  | Radiomics | 0.620 [0.547-0.693] |  | 293 | 300 | 0.586 | 0.656 | 0.543 | 0.695 |
| Logistic Regression | Clinical | 0.782 [0.716-0.838] | < 0.001 | 273 | 279 | 0.708 | 0.776 | 0.723 | 0.763 |
|  | Radiomics | 0.591 [0.510-0.675] |  | 310 | 316 | 0.605 | 0.647 | 0.489 | 0.746 |
| Random Forest | Clinical | 0.771 [0.706-0.833] | 0.002 | 270 | 277 | 0.687 | 0.770 | 0.723 | 0.737 |
|  | Radiomics | 0.613 [0.526-0.692] |  | 307 | 314 | 0.633 | 0.669 | 0.532 | 0.754 |
| XGBoost | Clinical | 0.766 [0.689-0.832] | 0.003 | 266 | 273 | 0.696 | 0.791 | 0.755 | 0.737 |
|  | Radiomics | 0.620 [0.523-0.696] |  | 308 | 315 | 0.596 | 0.667 | 0.564 | 0.695 |
| Hemorrhagic transformation | | | | | | | | | |
| GBM | Clinical | 0.634 [0.494-0.762] | 0.067 | 127 | 134 | 0.207 | 0.934 | 0.333 | 0.881 |
|  | Radiomics | 0.772 [0.667-0.866] |  | 122 | 129 | 0.147 | 0.990 | 0.944 | 0.490 |
| LightGBM | Clinical | 0.629 [0.404-0.761] | 0.031 | 128 | 135 | 0.118 | 0.974 | 0.889 | 0.381 |
|  | Radiomics | 0.787 [0.689-0.873] |  | 113 | 120 | 0.192 | 0.978 | 0.833 | 0.675 |
| Logistic Regression | Clinical | 0.656 [0.439-0.779] | 0.053 | 124 | 130 | 0.120 | 0.966 | 0.833 | 0.433 |
|  | Radiomics | 0.794 [0.679-0.889] |  | 113 | 120 | 0.289 | 0.970 | 0.722 | 0.835 |
| Random Forest | Clinical | 0.626 [0.407-0.759] | 0.003 | 126 | 133 | 0.175 | 0.948 | 0.556 | 0.758 |
|  | Radiomics | 0.833 [0.752-0.906] |  | 108 | 114 | 0.280 | 0.975 | 0.778 | 0.814 |
| XGBoost | Clinical | 0.589 [0.47-0.742] | 0.056 | 139 | 145 | 0.227 | 0.932 | 0.278 | 0.912 |
|  | Radiomics | 0.748 [0.651-0.838] |  | 126 | 133 | 0.160 | 0.991 | 0.944 | 0.541 |

Values for areas under the curves receiver operating characteristics are values [95% confidence intervals]. The *P*-values are used to compare the area under the receiver operating characteristic curves between the models using the radiomic features and the clinical variables on external validation.

AUC, area under the curve receiver operating characteristics AIC, Akaike information criterion; BIC, Bayesian information criterion; PPV, positive predictive value; NPV, negative predictive value.
